# Macrophage ferroptosis potentiates GCN2 deficiency induced pulmonary venous arterialization

**DOI:** 10.1101/2025.02.08.24318691

**Authors:** Jingyuan Zhang, Pei Mao, Tengfei Zhou, Bingqing Yue, Yaning Li, Yuanhua Qiu, Kejing Ying, Fudi Wang, Jingyu Chen, Jun Yang

**Affiliations:** Department of Physiology and Department of Cardiology of the Second Affiliated Hospital, Zhejiang University School of Medicine, Hangzhou 310058, China; State Key Laboratory of Transvascular Implantation Devices of the Second Affiliated Hospital, Zhejiang University School of Medicine, Hangzhou 310009, China; Center for Lung Transplantation of the Second Affiliated Hospital, Zhejiang University School of Medicine, Hangzhou 310052, China; Department of Pulmonary and Critical Care Medicine of Sir Run Run Shaw Hospital, Zhejiang University School of Medicine, Hangzhou 310016, China; Department of Public Health of Zhejiang University School of Medicine, Hangzhou 310058, China

## Abstract

Pulmonary veno-occlusive disease (PVOD) is a fatal disease characterized by the remodelling of pulmonary veins and haemosiderin accumulation in macrophages. Although GCN2 deficiency have been reported in PVOD patients, the underlying mechanism by which GCN2 deficiency affects the pulmonary venous cells and the surrounding cells, remains unclear. Here, we perform immunohistochemistry and scRNA-sequencing analyses to show that macrophages are the major population affected by GCN2 deficiency and ferroptosis pathway-related genes are upregulated in lung macrophages of PVOD patients. Treatment with the specific ferroptosis inhibitor ferrostatin-1 (Fer-1) reverses the changes in haemodynamic indices observed in *Eif2ak4*^K1488X/K1488X^ hypoxia mice and PVOD model rats. Furthermore, GCN2 deficiency increases HMOX1 and iron levels to facilitate ferroptosis in macrophages, and enhances arterial marker expression in venous endothelial cells (VECs). Specifically, spatial transcriptome analysis reveals increased expression of NRP1, KDR and EFNB2 through ETS1 in VECs from PVOD patients. Our findings suggest the potential of targeting macrophage ferroptosis as a therapeutic strategy for treating related vascular diseases, and of using NRP1/KDR/EFNB2 expression as a specific marker set for venous arterialization.

## Introduction

Pulmonary veno-occlusive disease (PVOD) is a heritable autosomal recessive disease characterized by the remodelling of pulmonary venules which leads to pulmonary hypertension (PH)^1, 2, 3^. PVOD typically has a poor prognosis, with a mean time from diagnosis to death or need for lung transplantation of less than 2 years^4, 5^. Biallelic *EIF2AK4* mutations have been found in all familial PVOD cases and in 9% of sporadic cases^4, 6^, with no sex-based differences reported. However, general control nonderepressible 2 (GCN2) deficiency is not only caused by biallelic *EIF2AK4* mutations, but also been found in cancer patients after treatment with alkylating chemotherapeutic agents, such as cyclophosphamide and mytomycin-C (MMC) or even higher tobacco exposure^7^. Because the mechanism underlying the effects of GCN2 deficiency on pulmonary venous remodelling is still unclear, lung transplantation remains the only available therapy for eligible patients^8^. Thus, evidence-based medical therapies for PVOD are needed^4, 9^.

GCN2, a serine-threonine kinase that is upregulated in response to various cellular stresses^10^, has been detected in pulmonary vessel walls and interstitial tissue, mostly in macrophages, in human lungs^11^, ^12^. Compared with gene-corrected isogenic controls, PVOD-iPSC-differentiated endothelial cells (ECs) exhibit excessive proliferation^13^. However, distinguishing changes in heterogeneous human endothelial cells (ECs), including artery, vein, and capillary ECs, can be challenging^14, 15, 16^, even though these EC types have distinct molecular signatures and different physiological functions depending on their formation and location within the vasculature^14^. Moreover, whether *EIF2AK4* mutations play specific roles in the VEC subtype fate switch and how the surrounding cells impact on VECs remain to be clarified.

In the lungs, macrophages are an abundant cell type that critically regulates normal homeostasis via the secretion of inflammatory factors and chemokines, which contribute to endothelium permeability and activation during the progression of pulmonary hypertension (PH) ^17^. As haemosiderin-laden macrophages in the lungs of PVOD patients is one of the main feather in PVOD, we hypothesize that *EIF2AK4* mutation alters the endothelium by affecting the iron metabolism of macrophages in the pulmonary vascular niche.

In this work we perform single-cell RNA transcriptome sequencing (scRNA-seq) on lung samples collected from PVOD patients and from mitomycin-C (MMC)-induced PVOD model rats (MMC rats) to identify the major affected cell types and key regulators involved in PVOD. Surprisingly, we identify arterial markers expressed in remodeled veins surrounded by haemosiderin-laden macrophages in the lungs of PVOD patients. Since the lack of an animal genetic model for *EIF2AK4* mutation-bearing PVOD is one reason for the limited understanding of this devastated disease^18, 19^, we generate *Eif2ak4*^K1488X/K1488X^ mice by knocking in the human disease-causing mutation and PVOD patient-induced pluripotent stem cells (iPSCs) to recapitulate the pathogenesis observed in the lungs of PVOD patients^20^. By combining scRNA-seq, spatial transcriptional sequencing and immunofluorescence staining, we demonstrate arterialization of the venous endothelium with markedly elevated NRP1/KDR/EFNB2 expression at vessel lesion sites. These findings suggest the important role of macrophage ferroptosis in arterialization of venous endothelium.

## Results

### Ferroptosis-related gene expression in lung macrophages is increased in PVOD patients with *EIF2AK4^mut^*

We sought to clarify the role of *EIF2AK4* mutation in the pathobiology of the pulmonary vasculature, especially the key factors that impact the development of occlusive venous lesion. All patients had pathogenic characteristics of advanced PVOD with human *EIF2AK4* mutation in two alleles with no sex bias (Fig. 1a). We carried out immunoblotting of GCN2 (the protein encoded by *EIF2AK4*) in lung tissues obtained from 5 control individuals and 5 patients bearing *EIF2AK4* mutations, which showed absence of GCN2 protein expression in the PVOD patients (Fig. 1b). Consistent with the immunoblotting results, immunostaining analysis revealed that GCN2 was highly expressed in macrophages from the lung tissues of control individuals, but not in PVOD patient lungs (Fig. 1c and Fig. S1a, b). In agreement with a previous study^21^, we also observed that the number of haemosiderin-laden macrophages was obviously increased in PVOD patients (Fig. 1d and Fig. S1c). The typical pathological features distinguishing PVOD from other forms of PH include pulmonary haemorrhage and accumulation of iron-containing haemosiderin deposits.

**Fig. 1.**
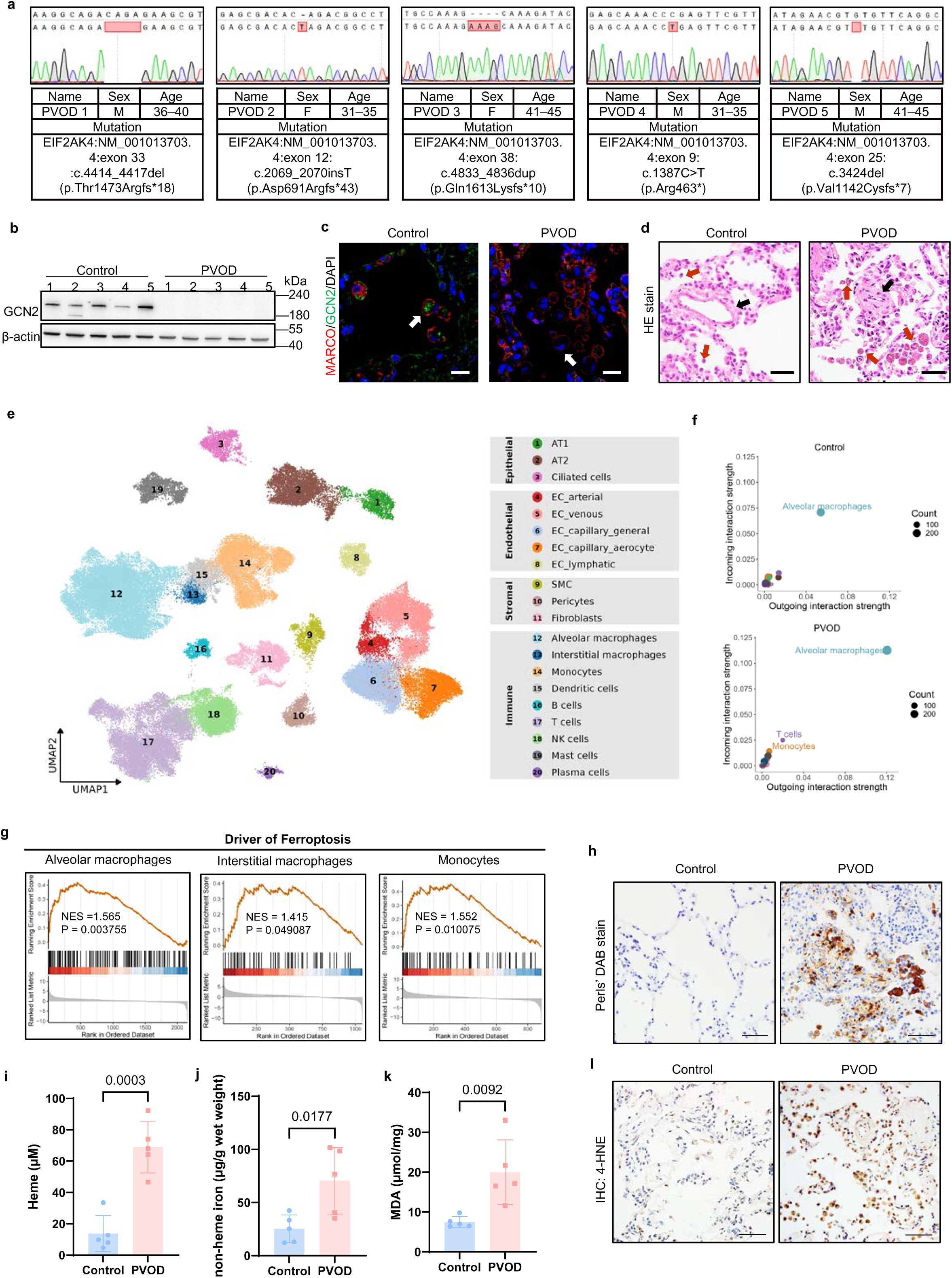
Ferroptosis-related gene expression in lung macrophages is increased in PVOD patients with *EIF2AK4*^mut^. **a** DNA sequencing analysis of tissues from pulmonary veno-occlusive disease (PVOD) patients in this study confirms EIF2AK4 mutations. M: Male, F: Female. **b** Western blot showing the expression levels of GCN2 in Control or PVOD patient lung tissues; n= 5 individuals. **c** Representative images of MARCO (red) and GCN2 (green) immunofluorescence and DAPI (blue) staining of lung tissues from Control and PVOD patients. The data are representative images, White arrow: macrophages. Scale bar=20 µm. n=6 microscope fields from 6 individuals with similar results. **d** Representative images of lung tissues stained with haematoxylin and eosin (H&E). Red arrow: macrophages; Black arrow: pulmonary vein. Scale bar=50 µm. Control, n=6 individuals; PVOD n=9 individuals with similar results. **e** UMAP projection showing the main cell type identified by integrated clustering analysis of scRNA-seq datasets from three Controls and three *EIF2AK4*^mut^ PVOD patients. **f** Bubble plot showing the incoming and outgoing interaction strengths for each subpopulation of immune cells in the Control and PVOD groups. The dot size represents the number of interactions. **g** GSEA of differentially expressed genes in alveolar macrophages, interstitial macrophages, and monocytes between the Control and PVOD groups. The ferroptosis-associated gene set was obtained from FerrDb. NES, normalized enrichment score. **h** Control and PVOD patient lung tissues were stained with Perls’ DAB to label ferric iron deposits. Scale bar=50 µm. Control, n=7 individuals; PVOD n=9 individuals with similar results. **i** Heme iron levels in the lungs of Control and PVOD patients were measured. n=5 individuals. **j** Non-heme iron levels in the lung were measured in Controls and PVOD patients. n=5 individuals. **k** The MDA content in the lungs of Control and PVOD patients was measured. n=5 individuals. **l** Representative images of IHC analysis of 4-HNE content in Control and PVOD patient lung tissues. Scale bar=50 µm. The data are presented as the means ± s.e.m. ; unpaired two-sided t test. Each dot represents an individual biological replicate, at least three independent experiments. *P* values are indicated in the figures. Source data are provided as a source data file.

Next we performed single-cell RNA sequencing (scRNA-seq) of explanted lung tissue samples from three patients with confirmed *EIF2AK4*^mut^ PVOD (obtained after lung transplantation) to define the involved cell types and their roles in this disease. We obtained high-quality transcriptomic data from 28,769 single cells after stringent segregation, and integrated this dataset with data from three control samples to perform cell type annotation on the basis of a reference dataset from the Human Lung Cell Atlas (HLCA) ^22, 23^. All the samples presented similar cell type compositions and proportions (Fig. 1e and Fig. S1d-f and Fig. S2). To identify the cell population that was most affected by *EIF2AK4*^mut^, we considered differences in cell–cell interactions to be potential sources of these phenotypic differences by comparing ligand-receptor interaction strengths among all the cell subtypes in the control individuals and PVOD patients. We found that the overall strength with which macrophages communicate with other cell types was significantly increased in PVOD patients (Fig. 1f).

Differential gene expression enrichment and pathway scoring analysis of macrophages including alveolar macrophages (AMs), interstitial macrophages (IMs), and monocytes, from controls and PVOD patients revealed that ferroptosis-associated gene expression was highly increased in PVOD patients (Fig. 1g and Fig. S1g). The degree of iron deposition, as determined by enhanced Perls’ DAB staining, progressively increased in PVOD patients (Fig. 1h, Figure S1h and S1i). Iron is required for the accumulation of lipid peroxidation products to induce ferroptosis^24^. We observed substantial lung iron accumulation (Fig. 1i, j) and lipid peroxidation, as evidenced by increased levels of MDA (Fig. 1k) and 4-HNE (Fig. 1l and Fig. S1j) expression in PVOD patients. These observations suggest that ferroptosis was induced in the lung macrophages of PVOD patients with *EIF2AK4*^mut^ .

### *Eif2ak4*^K1488X/K1488X^ mice as a model for inducing pulmonary venous remodelling in PVOD

To recapitulate the phenotype of genetic PVOD, we generated a knock-in mouse strain that bears the same disease-causing mutation (a premature stop codon) in exon 33 of the endogenous *EIF2AK4* locus as PVOD 1 and his family shown in Fig. 1 (Fig. 2a). Homozygous *Eif2ak4*^K1488X/K1488X^ mice presented normal birth rates, although GCN2 protein expression in the lungs was reduced in these mice (Fig. 2a). At 6 months of age, no detectable alterations in the total pulmonary vascular resistance index (TPVRI), the right ventricular systolic pressure (RVSP) and the extent of RVH were observed in *Eif2ak4*^K1488X/K1488X^ mice compared with wild-type (WT) mice (Fig. 2b-d). To induce a severe pulmonary vascular phenotype, we introduced hypoxia as a trigger for capillary contraction and endothelium leakage. After 6 weeks of hypoxia exposure, the TPVRI and RVSP values were significantly greater in the *Eif2ak4*^K1488X/K1488X^ mice than in the WT mice (Fig. 2b, c), reduced cardiac output (CO) was found in these *Eif2ak4*^K1488X/K1488X^ mice(Fig. S4b), but the extent of RVH was not significantly different between the two groups, even analysed with different sex (Fig. 2d).

**Fig. 2.**
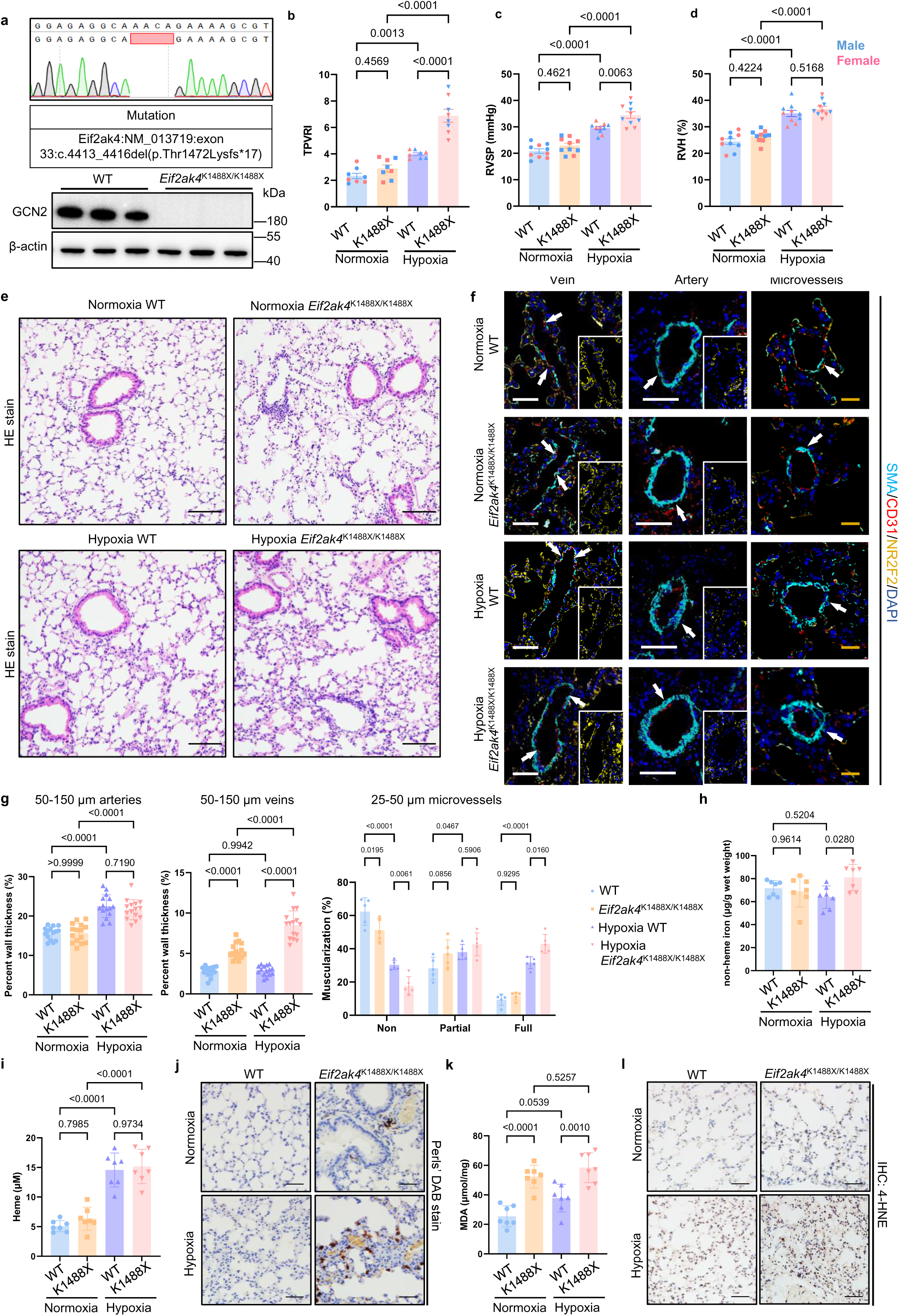
E*i*f2ak4K1488X^/K1488X^ mice as a model for inducing pulmonary venous remodelling in PVOD. **a** DNA sequencing analysis of *Eif2ak4*^K1488X/K1488X^ mice confirming the EIF2AK4 mutation. Western blot revealing decreased expression of GCN2 in *Eif2ak4*^K1488X/K1488X^ mouse lung tissues (n=3 mice/group). **b-d** Wild-type (WT) and *Eif2ak4*^K1488X/K1488X^ (K1488X) mice were subjected to normoxia or hypoxia for 6 weeks the total pulmonary vascular resistance index (TPVRI) value, right ventricular systolic pressure (RVSP) and right ventricular hypertrophy (RVH) in the model mice (b male:n=4 mice/group, female: n=4 mice /group; c and d male:n=5 mice/group; female: n=5 mice/group). TPVRI: Normoxia K1488X vs Hypoxia K1488X, *p*=3.5×10^-9^; Hypoxia WT vs K1488X, *p*=7×10^-7^. RVSP: Normoxia K1488X vs Hypoxia K1488X, *p*=2×10^-8^; Normoxia WT vs Hypoxia WT, *p*=4.6×10^-7^. RVH: Normoxia K1488X vs Hypoxia K1488X, *p*=7×10^-8^; Normoxia WT vs Hypoxia WT, *p*=5×10^-8^. **e** Representative images of H&E-stained samples from the model mice. Scale bar=100 µm. **f** Representative images of α-SMA (cyan), CD31(red), NR2F2 (yellow) and DAPI (blue) immunofluorescence staining of veins (with NR2F2 and DAPI co-staining), arteries and micrevessels from WT and *Eif2ak4*^K1488X/K1488X^ mice under nomaxia and hypoxia. White scale bar=50 µm. Yellow scale bar=20 µm. White arrow: vessel. **g** Percent wall thickness of pulmonary arteries (n=15 vessels of 7 mice/group), medial thickness of pulmonary veins (n=15 vessels of 7 mice/group), and muscularization (%) of microvessels (n=5 mice/group) in the model mice. Arteries: Normoxia K1488X vs Hypoxia K1488X, *p*=6×10^-8^; Normoxia WT vs Hypoxia WT, *p*=2×10^-9^. Veins: Normoxia K1488X vs Hypoxia K1488X, *p*=6×10^-9^; Normoxia WT vs K1488X, *p*=1×10^-6^; Hypoxia WT vs K1488X, *p*=1×10^-12^. Microvessels: Non WT vs Hypoxia WT, *p*=9×10^-11^; Full WT vs Hypoxia WT, *p*=1×10^-6^. **h** Non-heme iron levels in the lungs of the mice were measured. n=7 mice/group. **i** Heme iron levels in the lungs of the mice were measured. n=7 mice/group. Normoxia K1488X vs Hypoxia K1488X, *p*=2.9×10^-5^; Normoxia WT vs Hypoxia WT, *p*=1.1×10^-5^. **j** Ferric iron deposits in the model mice were stained with Perls’ DAB. Scale bar=50 µm. **k** MDA content in the lungs of the mice was measured. Normoxia WT vs K1488X, *p*=5×10^-5^. **l** Representative images of 4-HNE staining in mice. Scale bar=50 µm. The data are presented as the means ± s.e.m.; two-way ANOVA with Tukey’s multiple comparison test. Each dot represents an individual biological replicate, at least three independent experiments. *P* values are indicated in the figures. Source data are provided as a source data file.

Histopathological analysis revealed the substantial infiltration of inflammatory cells around the pulmonary vein in *Eif2ak4*^K1488X/K1488X^ mice under hypoxia (Fig. 2e). Morphological analysis of vessels via immunofluorescent staining revealed wall thickening within the pulmonary arterioles in WT and *Eif2ak4*^K1488X/K1488X^ mice under hypoxia; strikingly, the muscularization of venules surrounded by inflammatory cells, indicating venous remodelling, was specifically detected in *Eif2ak4*^K1488X/K1488X^ mice with or without hypoxia (Fig. 2f, g and Fig. S4g). We found irreversible PVOD phenotype in *Eif2ak4*^K1488X/K1488X^ mice compared with WT after reoxgenation (Fig. S3). Furthermore, GCN2-deficient bone marrow transplanted (BMT) mice had significantly elevated TPVRI, RVSP and worse right heart function than WT BMT mice under hypoxia (Fig. S5).

We also detected significantly increased iron levels in whole-mouse-lung homogenates from *Eif2ak4K*^1488X/K1488X^ mice under hypoxia (Fig. 2h). Interestingly, heme-iron levels were significantly increased in hypoxic mice, but did not elevated in *Eif2ak4K*^1488X/K1488X^ mice compared with control mice under hypoxia (Fig. 2i). Consistent with the heme iron levels, we found no significant difference in endothelium permeability between WT and *Eif2ak4K*^1488X/K1488X^ mice under hypoxia (Fig. S4c). This finding suggested that blood cells that leak through the endothelium are not the source of the elevated iron observed in mutant mouse lungs, another possibility could be that iron recycling is defective in mutant macrophages, leading to iron accumulation. Moreover, Perls’ DAB staining revealed that macrophages in the lungs of *Eif2ak4K*^1488X/K1488X^ mice presented greater levels of iron deposition than those in the lungs of WT mice under hypoxia did (Fig. 2j and Fig. S4d). To confirm the occurrence of macrophage ferroptosis, we then measured lipid peroxidation in the lung tissue. Significantly elevated MDA levels (Fig. 2k) and enhanced 4-HNE staining of macrophages (Fig. 2l and Fig. S4e) were also observed in the lungs of *Eif2ak4K*^1488X/K1488X^ mice under hypoxia. Moreover, we examined iron accumulation in other organs, including the kidney, liver, spleen and heart. Increased iron deposition was detected in the liver and spleen but not in the kidney or heart (Fig. S4f). Taken together, these findings suggest that macrophages bearing the *Eif2ak4* mutation were refractory to iron recycling, which contributed to pulmonary venous remodelling in the mice.

### Fer-1 reverses PVOD in *Eif2ak4*^K1488X/K1488X^ hypoxia-induced mice and MMC rats

To confirm the importance of macrophage ferroptosis in PVOD development, we examined the effects of the ferroptosis inhibitor ferrostatin-1 (Fer-1) in the established PVOD mouse model. *Eif2ak4*^K1488X/K1488X^ mice were reared under hypoxic or normoxic conditions for 6 weeks and treated with vehicle or Fer-1 either from the beginning of the hypoxia period (prevention protocol) or beginning on day 21 of hypoxia exposure (reversal protocol) (Fig. 3a). We observed that Fer-1 treatment significantly reduced the TPVRI, RVSP and RVH in mice treated with both protocols, even analysed with different sex (Fig. 3b). Fer-1 reversed the histopathological phenotype of PVOD, including reductions in the venous wall thickness and inflammatory cells infiltration (Fig. 3c and Fig. S4h). Morphometric analysis and quantification of α-SMA staining revealed reduced thickening of the pulmonary artery and venous wall and muscularization of microvessels in the mice subjected to both protocols (Fig. 3d, e). Wheras Fer-1 cannot reverse pulmonary hypertension in hypoxic WT mice (Fig. S7). Together, these findings confirmed that ferroptosis inhibition reversed the PVOD phenotype in *Eif2ak4*^K1488X/K1488X^ hypoxia mice.

**Fig. 3.**
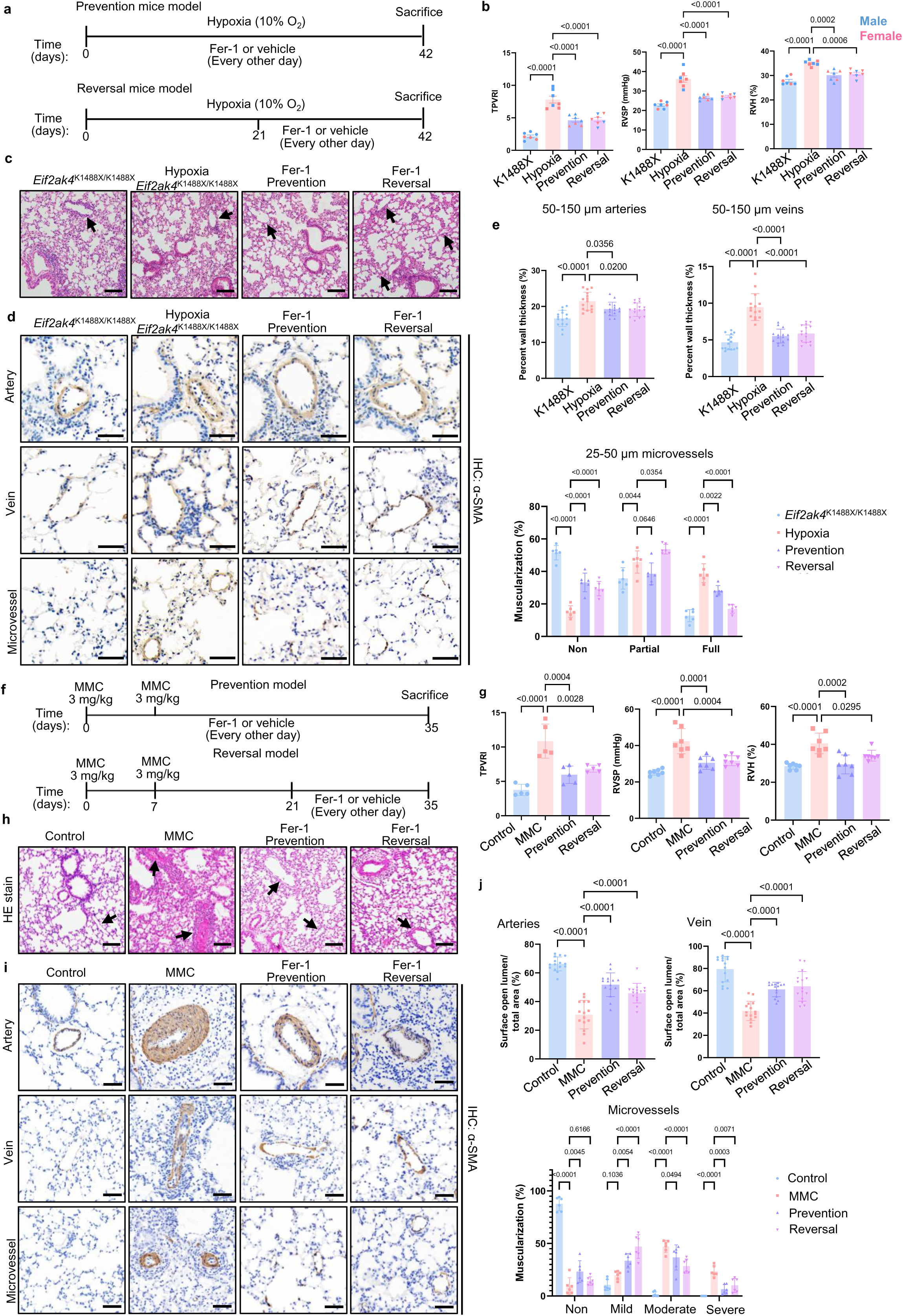
Fer-1 reverses PVOD in *Eif2ak4*^K1488X/K1488X^ hypoxia-induced mice and MMC rats. **a** Schematic diagram of the experimental design. *Eif2ak4*^K1488X/K1488X^ ^mice^ (K1488X) were subjected to hypoxic or normoxic conditions for 42 days and treated with vehicle or Ferrostatin-1 (Fer-1) from the start of the hypoxia treatment (prevention protocol) or beginning at 21 days (reversal protocol). **b** Assessment of TPVRI, RVSP and RVH in the mice (TPVRI male:n=4,4,4,3 mice/group, female: n=3,3,3,4 mice/group; RVSP male:n=4,3,4,3 mice/group, female: n=3,4,3,4 mice/group; RVH male:n=4,4,4,3 mice/group, female: n=3,3,3,4 mice/group). TPVRI: K1488X vs Hypoxia, *p*=3.9×10^-9^; Hypoxia vs Prevention, *p*=1.8×10^-5^; Hypoxia vs Reversal, *p*=2×10^-5^. RVSP: K1488X vs Hypoxia, *p*=6×10^-8^; Hypoxia vs Prevention, *p*=9×10^-6^; Hypoxia vs Reversal, *p*=2×10^-5^. RVH: K1488X vs Hypoxia, *p*=9.7×10^-7^. **c** Representative images of H&E-stained lung sections from the mice described in a. Pulmonary veins (arrows), scale bar=100 µm. The experiment was repeated independently 5 times with similar results. **d** Representative images of α-SMA staining in lung sections from the mice described in a. Scale bar=50 µm. **e** Percent wall thickness of pulmonary arteries (n=15 vessels of 7 mice /group), medial thickness of pulmonary veins (n=15 vessels of 7 mice/group), and muscularization (%) of microvessels (n=6 mice/group) in the mice described in a. Arteries: K1488X vs Hypoxia, *p*=5.8×10^-7^. Veins: K1488X vs Hypoxia, *p*=7.5×10^-12^; Hypoxia vs Prevention, *p*=1×10^-10^; Hypoxia vs Reversal, *p*=1.7×10^-9^. Microvessels: Non K1488X vs Hypoxia, *p*=2×10^-11^; Hypoxia vs Prevention, *p*=2×10^-7^; Hypoxia vs Reversal, *p*=3.9×10^-5^. Full K1488X vs Hypoxia, *p*=2×10^-11^; Hypoxia vs Reversal, *p*= 9×10^-10^. **f** Schematic diagram of the experimental design. The rats were given 3 mg/kg mitomycin-C (MMC) (once per week for 2 weeks, i.p.) or vehicle for 35 days and Fer-1 or saline from the start of the MMC treatment (prevention protocol) or beginning at 21 days (reversal protocol). **g** Assessment of TPVRI, RVSP and RVH in the rats (n=7 rats/group) described in f. TPVRI: Control vs MMC, *p*=5.9×10^-6^; RVSP: Control vs MMC, *p*=4×10^-7^; RVH: Control vs MMC, *p*=5×10^-5^. **h** Representative images of H&E-stained lung sections from the rats described in f. Pulmonary veins (arrows), scale bar=100 µm. The experiment was repeated independently 5 times with similar results. **i** Representative images of α-SMA staining in the pulmonary arteries, veins and microvessels of the rats described in f. Scale bar=50 µm. **j** Surface open lumen/total area (%) of pulmonary arteries and veins (n=15 vessels of 5 rats/group) and muscularization (%) of microvessels (n=6 rats/group) in the rats described in f. Arteries: Control vs MMC, *p*=7×10^-12^; MMC vs Prevention, *p*=4×10^-9^; MMC vs Reversal, *p*=1×10^-5^. Veins: Control vs MMC, *p*=8×10^-12^; MMC vs Prevention, *p*= 2×10^-5^; MMC vs Reversal, *p*=1×10^-6^. Microvessels: Non Control vs MMC, *p*<1×10^-15^; Mild MMC vs Reversal, *p*=9.6×10^-9^; Moderate Control vs MMC, *p*<1×10^-15^, MMC vs Reversal, *p*=6×10^-5^; Severe Control vs MMC, *p*=5.7×10^-7^. The data are presented as the means ± s.e.m.; one-way ANOVA with Tukey’s multiple comparison test. Each dot represents an individual biological replicate, at least three independent experiments. *P* values are indicated in the figures. Source data are provided as a source data file.

To investigate whether macrophage ferroptosis, which was observed in the genetic form of PVOD, also occurs in sporadic PVOD. We performed scRNA-seq on whole lung tissues from control rats and MMC-treated rats (female, which are more sensitive to MMC treatment^25^), a non-genetic model of disease in which reduction of GCN2 expression is a central feature (Fig. S8a, b). These MMC rats exhibited typical phenotypes of PVOD, including reduced pulmonary artery and venous lumen areas and capillary haematomas (Fig. S8e). A total of 24,156 cells from 4 samples (from 2 control rats and 2 MMC rats) were included after quality control (Fig. S9). Combined Seurat analysis provided detailed classification of immune cells (Fig. S9c). Enrichment scores for iron accumulation signatures (IAS)^26^ and ferroptosis gene sets^27^ were calculated, revealing significant elevation in macrophages, within the MMC group (Fig. S8c, d). Iron levels in whole-lung homogenates were significantly increased in MMC rats (Fig. S8f, g). Macrophages had greater levels of iron deposition in the MMC than in control rat lungs (Fig. S8h, i). Significantly elevated lipid peroxidation were also observed in the MMC rats (Fig. S8j-l). Overall, consistent with our observations showed above, ferroptotic macrophages and accumulated iron in interstitium may induce the PVOD phenotype in MMC rats.

We examined the effects of Fer-1 in MMC rats^28^. The rats were given 3 mg/kg MMC (once per week for 2 weeks, i.p.) for 5 weeks, with or without Fer-1 administered either from the beginning of MMC treatment (prevention protocol) or starting on day 21 of MMC treatment (reversal protocol) (Fig. 3f). Fer-1 reduced the TPVRI, RVSP and ameliorated RVH in rats treated with both prevention and reversal protocols (Fig. 3g). Similarly, Fer-1 prevented or rescued the histopathological phenotype of PVOD, reversing the pulmonary artery and venous lumen areas and the incidence of capillary haematomas, in the treatment groups (Fig. 3h). Morphometric analysis revealed reduced initial narrowing within pulmonary arteries and veins, and quantification of α-SMA staining indicated alleviation of the muscularization microvessels in rats treated with both prevention and reversal protocols (Fig. 3i, j).

### GCN2 deficiency enhances HMOX1 expression and promotes ferroptosis in macrophages

To identify the downstream mediator of GCN2, we performed differentially expressed genes (DEGs) analysis of scRNA-seq data from macrophages and determined that *HMOX1* was among the top DEGs (Fig. 4a). The protein encoded by *HMOX1*, heme oxygenase 1 (HMOX1), catalyses the rate-limiting step in heme degradation to release iron, and an aberrant elevation in HMOX1 levels may initiate ferroptosis under pathological conditions^29^. We performed Western blot analysis of HMOX1 and the ferroptosis markers FTL, FTH and TFRC in all the PVOD patient lung tissue samples and observed significantly elevated protein expression compared with that in the control samples (Fig. 4b, c). The expression of all of these factors was significantly increased in whole-lung tissue homogenates from MMC rats (Fig. 4e, f) and *Eif2ak4*^K1488X/K1488X^ hypoxia mice (Fig. 4h, i) and in *Eif2ak4*^K1488X/K1488X^ bone marrow-derived macrophages (BMDMs) (Fig. 4j, k). Increased HMOX1 expression was also confirmed in GCN2 knockout HT1080 cells in the presence of additional iron (Fig. S10a, b). Furthermore, the immunofluorescence results confirmed that HMOX1 was highly expressed in macrophages from PVOD patients and MMC rats (Fig. 4d, g).

**Fig. 4.**
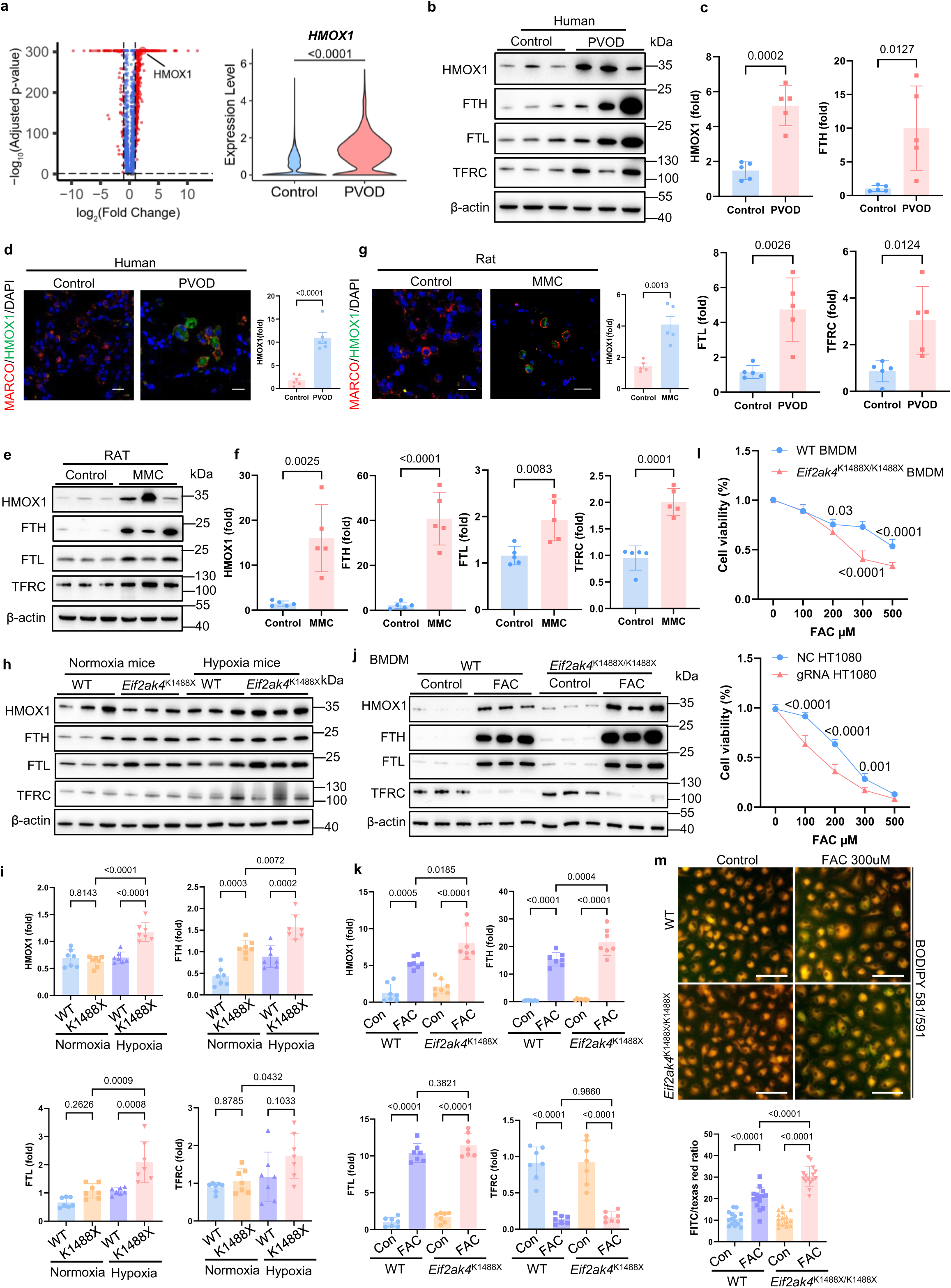
GCN2 deficiency enhances HMOX1 expression and promotes ferroptosis in macrophages. **a** Volcano plot depicting the differentially expressed genes in alveolar macrophages from PVOD patients and Controls. The violin plot highlights HMOX1 expression, identified as significantly different. *P* values were determined via two-sided Wilcoxon rank-sum test with Bonferroni correction for multiple testing. *p*=1.42e-686. **b, c** Western blot and quantification of HMOX1, FTH, FTL, and TFRC in Control and PVOD lung tissues. Data are presented as the means ± s.e.m. (n=5 individuals); unpaired two-sided t test. **d** Representative images and quantification of MARCO(red) and HMOX1(green) immunofluorescence and DAPI (blue) staining of lung tissues from Control and PVOD patients. Scale bar=20 µm. Data are presented as the means ± s.e.m. (n=6 individuals); unpaired two-sided t test. HMOX1, *p*=3.5×10^-5^. **e-f** Western blot of HMOX1, FTH, FTL, and TFRC in Control and MMC rat lungs. Data are presented as the means ± s.e.m. (n=5 rats/group); unpaired two-sided t test. FTH, *p*=8.39×10^-5^. **g** Representative images and quantification of MARCO(red) and HMOX1(green) immunofluorescence and DAPI (blue) staining of lung tissues from Control and MMC rats. Scale bar=20 µm. Data are presented as the means ± s.e.m. (n=5 rats/group); unpaired two-sided t test. **h, i** WT and *Eif2ak4*^K1488X/K1488X^ mice were subjected to hypoxia for 6 weeks. Western blot of HMOX1, FTH, FTL, and TFRC in mouse lungs. Data are presented as the means ± s.e.m. (n=7 mice/group); two-way ANOVA with Tukey’s multiple comparison test. HMOX1, Hypoxia WT vs K1488X, *p*=1.7×10^-5^; Normoxia K1488X vs Hypoxia K1488X, *p*=2.3×10^-6^. **j, k** Western blot of HMOX1, FTH, FTL and TFRC in WT or *Eif2ak4*^K1488X/K1488X^ BMDMs treated with or without 300 µM FAC for 48 h. Data are presented as the means ± s.e.m. (n=7 mice/group); two-way ANOVA with Tukey’s multiple comparison test. HMOX1, K1488X Con vs FAC, *p*=5×10^-6^; FTH, WT Con vs FAC, *p*=2×10^-9^, K1488X Con vs FAC, *p*=9.8×10^-12^; FTL, WT Con vs FAC, *p*=9.7×10^-11^, K1488X Con vs FAC, *p*=5×10^-11^; TFRC, WT Con vs FAC, *p*=2×10^-6^, K1488X Con vs FAC, *p*=2.7×10^-6^. **l** Cell viability was measured in WT and *Eif2ak4*^K1488X/K1488X^ BMDMs treated with the indicated concentrations of FAC for 48 h and in normal control (NC) and GCN2^-/-^ (gRNA) HT1080 cells treated with the indicated concentrations of FAC for 48 h. Data are presented as the means ± s.e.m. (n = 6 replicates); unpaired two-sided t tests. BMDM, FAC 300 µM, *p*=1.3×10^-6^, FAC 500 µM, *p*=8.3×10^-5^; HT1080, FAC 100 µM, *p*=3×10^-5^, FAC 200 µM, *p*=1.1×10^-5^. **m** Lipid peroxidation in BMDMs treated with FAC for 12 h, assayed using BODIPY 581/591 dye. Representative microscopy images shown. Scale bar=50 µm. The data are presented as the means ± s.e.m. (n=15 microscope fields from 5 mice); two-way ANOVA with Tukey’s multiple comparison test. WT Con vs FAC, *p*=9×10^-8^, K1488X Con vs FAC, *p*=1×10^-12^, FAC WT vs K1488X, *p*=1×10^- 8^. Each dot represents an individual biological replicate, at least three independent experiments. *P* values are indicated in the figures. Source data are provided as a source data file.

These findings suggest that GCN2 deficiency renders macrophages more susceptible to ferroptosis. To investigate this hypothesis, we evaluated the effects of the ferroptosis activators ferric citrate (FAC) in BMDMs obtained from WT and *Eif2ak4*^K1488X/K1488X^ mice. Furthermore, we generated *EIF2AK4* knockout HT1080 cells (a cell line sensitive to ferroptosis) via CRISPR/Cas9 targeted editing to examine the effects of ferroptosis activation in these cells and in control cells. Treatment with FAC reduced cell viability in a dose-dependent manner in all the cells examined, and this ferroptosis-activating effect was stronger in the mutant cells than in the normal control cells (Fig. 4l). BODIPY 581/591 staining revealed greater lipid peroxidation in response to FAC exposure in *Eif2ak4*^K1488X/K1488X^ BMDMs than in WT BMDMs (Fig. 4m). Moreover, we observed a significant increase in intracellular iron deposition in GCN2-deficient BMDMs and HT1080 cells following iron treatment (Fig. S10 c-f). However, the GSH/GSSG ratio, an indicator of redox status, did not significantly change in human lung tissue or iron-treated cells (Fig. S10g, h). Also, the expression of other ferroptosis-related factors, including ACSL4, TXNRD1 and GPX4 were not significantly changed which excluded the possibility of the involvement of these factors (Fig. S11). These findings suggested that GCN2 deficiency sensitized macrophages to ferroptosis induction via increased HMOX1 expression, especially under iron stimulation.

Previous studies have shown that GCN2 deficiency leads to recruitment of the transcription factor NRF2 to the *HMOX1* gene, resulting in elevated HMOX1 expression^30, 31^. Therefore, we analysed the expression of NRF2 in BMDMs and HT1080 cells by immunoblotting. Consistent with previous findings, GCN2-deficient cells presented a significant increase in NRF2 expression under iron stimulation (Fig. S12a-d). To examine whether HMOX1 could exaggerate the phenotype in our model mice, we challenged *Eif2ak4*^K1488X/K1488X^ mice with the HMOX1 activator hemin. The stimulation of *Eif2ak4*^K1488X/K1488X^ mice with hemin developed PH phenotypes within 6 weeks (Fig. S12e). However, the treatment of PVOD model mice and model rats with either an HMOX1 activator, hemin, or an HMOX1 inhibitor, Znpp, did not significantly promote or inhibit PVOD progression (Fig. S12f, g). Collectively, these findings indicate that elevated levels of HMOX1/FTL/FTH increase the susceptibility of GCN2-deficient macrophages to ferroptosis, but targeting HMOX1 alone is not desirable for the treatment of PVOD.

### GCN2 deficiency enhances VEC arterialization and smooth muscle cell recruitment in the presence of iron

To investigate the effects of macrophages on the pulmonary vasculature, we next examined the DEGs in *EIF2AK4*^mut^ VECs in the lungs of PVOD patients. ScRNA-seq revealed that genes related to arterial development (*NRP1, KDR,* and *EFNB2),* the arterial specification-related genes *NOTCH4* and *JAG2*, and the secretory marker *CXCL12* were highly upregulated in PVOD VECs compared with control VECs, whereas the venous related genes *TEK*, *ACKR1* and *ENG* presented reduced expression (Fig. 5a). When we specifically extracted the VECs subpopulation for pseudotime trajectory analysis, we found that VECs from PVOD patients were predominantly distributed in the high pseudotime regions (Fig. S2d, e). This finding supports our hypothesis regarding venous fate transition in PVOD pathogenesis, providing further evidence for the venous-to-arterial remodelling process at the single-cell level. These finding suggest that venous arterialization may occur in the lungs of PVOD patients. Venous arterialization occurs naturally during certain developmental processes, such as the initial derivation of coronary arteries from venous VECs in mice. EFNB2 (Ephrin-B2) signaling contributes to arterial specification via KDR activity^32^. Moreover, NRP1 serves as a co-receptor for VEGF-A and plays a pivotal role in arteriogenesis by facilitating KDR endocytosis and trafficking^33^. We employed the CRISPR/Cas9 system to knock in the mutated *EIF2AK4* gene (GCN2^-/-^) in human umbilical vein endothelial cells (hUVECs), the high efficiency of this process was confirmed (Fig. S13a). Treatment of GCN2^-/-^ hUVECs with macrophage-conditioned medium led to the upregulation of arterialization-related genes (Fig. S13b).

**Fig. 5.**
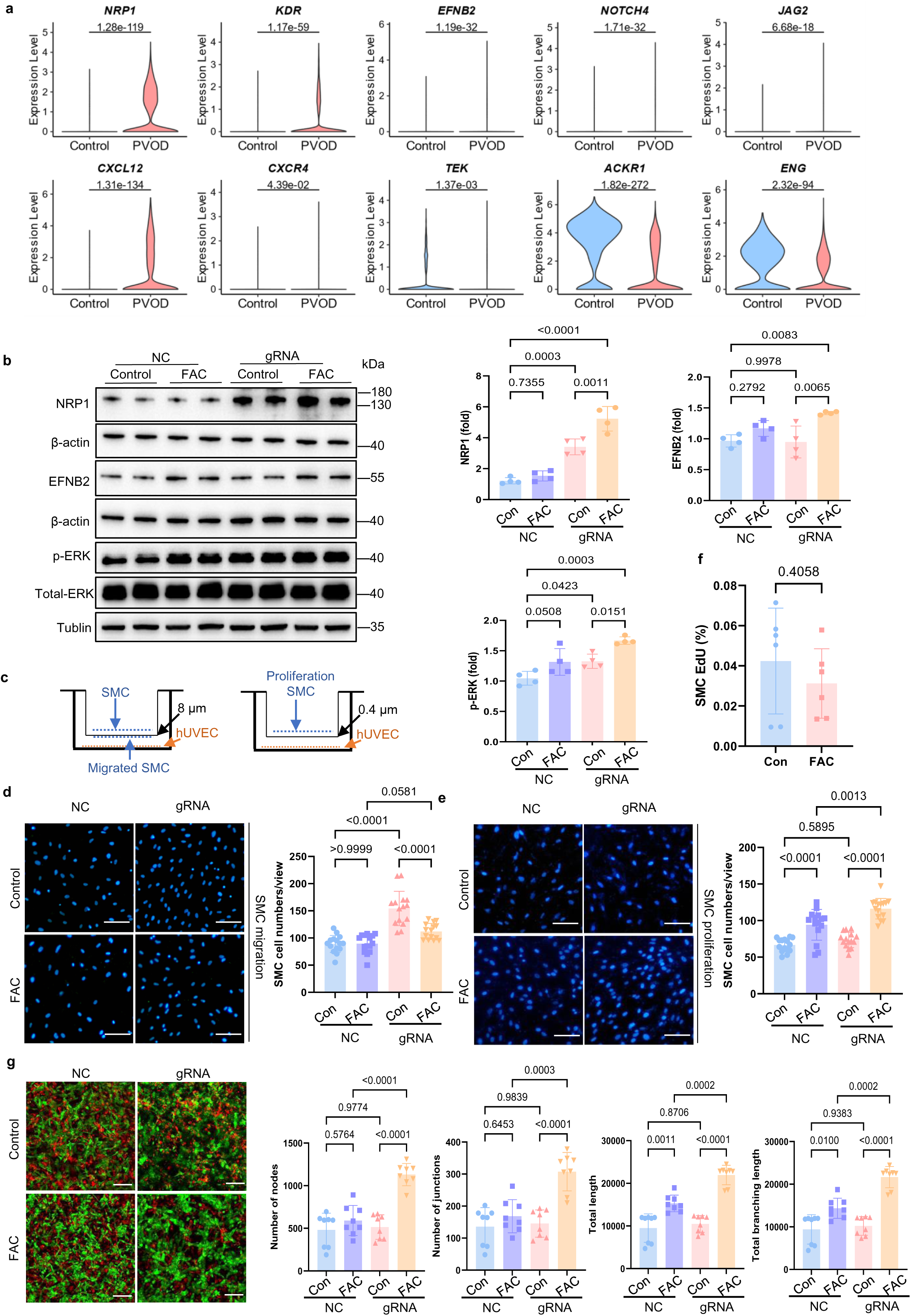
GCN2 deficiency enhances arterial markers expression in VECs and smooth muscle cell recruitment in the presence of iron. **a** Violin plots highlighting the differentially expressed genes associated with arterial and venous identity in venous endothelial cells from human scRNA-seq data. *P* values were determined via two-sided Wilcoxon rank-sum test and marked on each panel. **b** Western blot of NRP1, EFNB2, p- ERK in NC or GCN2^-/-^ (gRNA) hUVECs treated with/without 660 µM FAC for 72 h. Data are presented as the means ± s.e.m. (n=4 replicates); two-way ANOVA with Tukey’s multiple comparison test. NRP1, NC Con vs gRNA FAC, *p*=2×10^-6^. **c** Schematic of the Transwell coculture model. NC or GCN2^-/-^ hUVECs were seeded in the bottom compartment and treated with/without 660 µM FAC for 72 h, with SMCs seeded on top for 24 h. An 8-µm pore membrane facilitates migration; a 0.4-µm membrane tests proliferation. **d** Images and quantification of DAPI-stained SMCs cocultured with NC or GCN2^-/-^ (gRNA) hUVECs in an 8-μm pore size chamber in FAC-treated medium indicating the migration ability of SMCs. Scale bar=100 µm. Data are presented as the means ± s.e.m. (n=15 microscope fields from 3 independent experiments); two-way ANOVA with Tukey’s multiple comparison test. Con NC vs Con gRNA, *p*=3×10^-10^, gRNA Con vs FAC, *p*=1×10^-5^. **e** Images and quantification of DAPI-stained SMCs cocultured with NC or GCN2^-/-^ hUVECs in a 0.4-μm pore size chamber in FAC-treated medium revealing the proliferative ability of the SMCs. Scale bar=100 µm. The data are presented as the means ± s.e.m. (n=15 microscope fields from 3 independent experiments); two-way ANOVA with Tukey’s multiple comparison test. NC Con vs FAC, *p*=5×10^-5^, gRNA Con vs FAC, *p*=6×10^-8^. **f** Ratio of EdU-positive SMCs after treatment ± 660 µM FAC for 24 h. Data are presented as the means ± s.e.m. (n=6 replicates); unpaired two-sided t test. **g** Representative images of hUVECs (green) cocultured with SMCs (PKH26, red) for 4 days. Scale bar=100 µm. Quantification of total network area, total cord length, and branch points. Data are presented as the means ± s.e.m. (n=8 microscope fields from 4 independent experiments); two-way ANOVA with Tukey’s multiple comparison test. Number of nodes, NC FAC vs gRNA FAC, *p*=2×10^-5^, gRNA Con vs FAC, *p*=2×10^-6^; Number of junctions, gRNA Con vs FAC, *p*=4.5×10^-5^; Total length, gRNA Con vs FAC, *p*=8×10^-8^; Total branching length, gRNA Con vs FAC, *p*=3×10^-7^. Each dot represents an individual biological replicate, at least three independent experiments. *P* values are indicated in the figures. Source data are provided as a source data file.

Screening for secretory factors in human lung macrophages via scRNA-seq revealed that *GDF15*, *TNF* and *CCL3* were highly expressed (Fig. S13c, d). The elevated expression of GDF15, TNF and CCL3 was validated by ELISA of homogenized human lung tissue and supernatants collected from iron-treated *Eif2ak4*^K1488X/K1488X^ BMDMs (Fig. S13e). However, the treatment of GCN2^-/-^ hUVECs with these factors did not induce arterialization (Fig. S13f). In addition to the high levels of inflammatory factors in ferroptotic macrophage supernatants, iron can induce fibrosis in vascular disease, as recently reported^26, 34^. In this study, we detected increased expression of arterialization- related genes in iron-treated GCN2^-/-^ hUVECs (Fig. 5b and Fig. S14). To examine the effect of iron on the vasculature, we conducted a Transwell study of human pulmonary smooth muscle cells (SMCs) with GCN2^-/-^ hUVECs plated at the bottom well of the Transwell plate and observed an increase in SMCs migration (Fig. 5c, d). FAC treatment on hUVECs further increased the proliferation of SMCs (Fig. 5c, e), which was not affected by the addition of iron to the culture medium (Fig. 5f). Furthermore, we established a 3D hUVECs–SMCs coculture assay to confirm the effect of iron on endothelial network formation(Fig. 5g). Compared with the control culture conditions, GCN2^-/-^ hUVEC cocultures resulted an increase in the numbers of SMCs that wrapping around the ECs to provide physical support and for tube formation. Compared with control cultures, FAC-treated GCN2^-/-^ hUVEC cocultures presented increases in the total endothelial network area, total cord length and number of branch points. We also examined human pulmonary artery endothelial cells (hPAECs) or human pulmonary microvascular cells (hPMECs) treated with iron. There were no significant changes in gene expression in the hPAECs (Fig. S15a-c), whereas GCN2 deficiency in hPMECs resulted in increased *KDR*, *NOTCH4*, and *JAG2* expression, as well as the increased expression of *Ki67* and *PCNA*, which are associated with vascular malformation (Fig. S15d-f).

To determine the direct effects of iron on the GCN2-deficient pulmonary vasculature in vivo, we intratracheally administered FAC to *Eif2ak4*^K1488X/K1488X^ mice. HE staining of the lungs revealed the persistent infiltration of inflammatory cells and deposition of haemosiderin after 14 days of intrapulmonary iron administration (Fig. S6a). By day 35, the infiltration of inflammatory cells had decreased, but significant thickening of the blood vessel walls was observed. Iron staining of the lung tissue revealed abundant iron deposition from days 14 to 21, particularly in macrophages. However, iron deposition gradually decreased after 35 days of iron administration (Fig. S6b). Notably, the RVSP and the extent of RVH in the mice significantly increased after 14 days of iron administration, but did not decrease after 35 days, despite the decrease in iron deposition (Fig. S6c, d). Although there was no significant change in the thickness of the medial layer in the pulmonary arteries on day 35, significantly increased medial thickness in the pulmonary veins and microvessel muscularization was observed (Fig. S6e-g). In summary, GCN2 deficiency enhanced arterial marker gene expression in VECs and iron treatment promoted venous medial thickening in vitro and in vivo.

### Enhanced NRP1/KDR/EFNB2 expression as a marker set of the arterialization of pulmonary VECs

To investigate the spatial distribution of venous arterialization in PVOD, we performed 10X Visium spatial transcriptomics on lung tissues from a PVOD patient and two healthy controls (Fig. 6a). Integration of samples using STAligner^35^ followed by Leiden clustering identified nine distinct clusters that were categorized into four tissue regions: Alveoli, Bronchi, Vessel, and Unspecified areas based on marker gene expression and histological features (Fig. 6b and Fig. S16a-e).

**Fig. 6.**
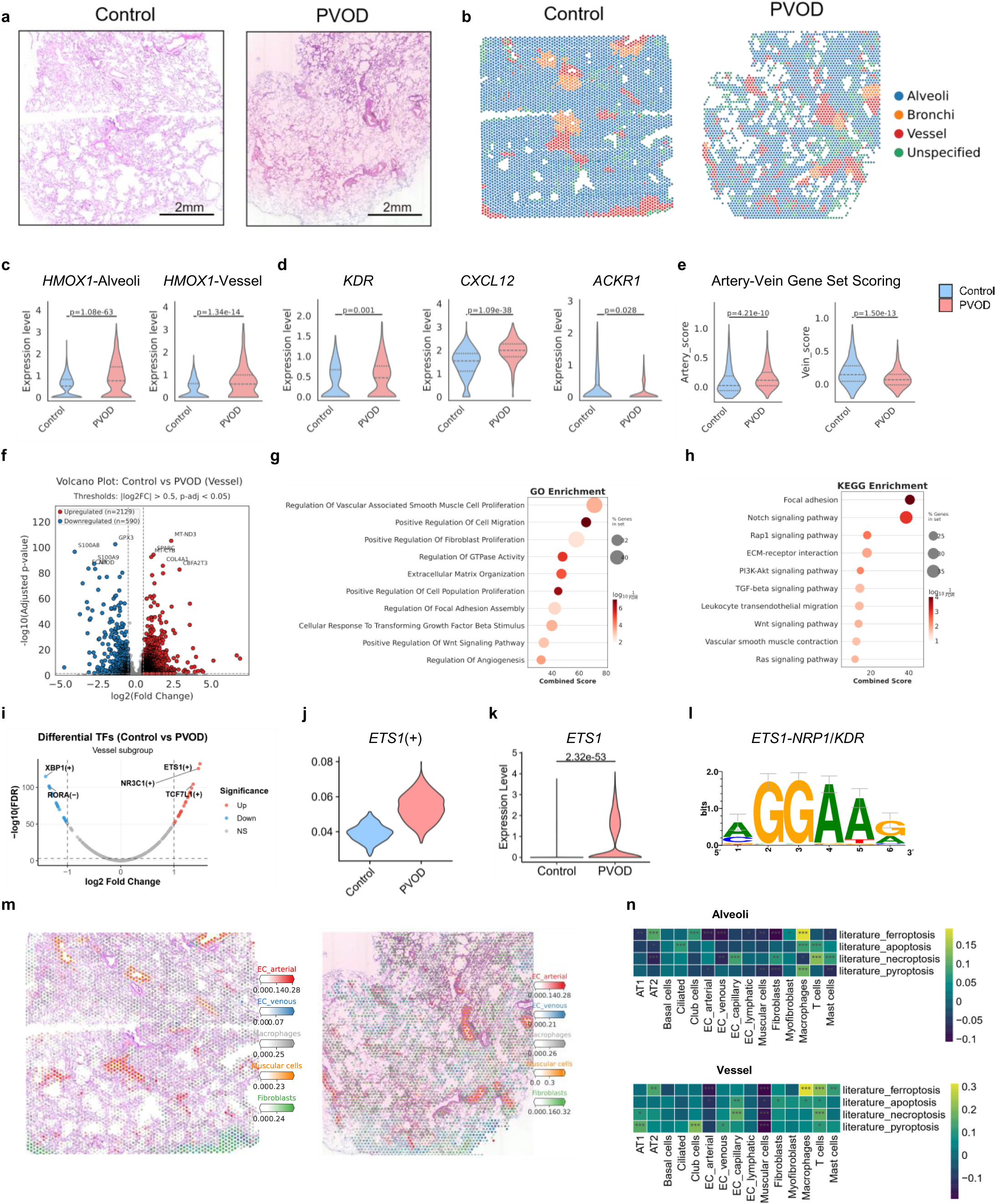
Spatial transcriptomics reveals enhanced venous arterialization and ETS1-mediated gene regulation in PVOD lung vessels. **a** H&E-stained images of 10X Visium spatial transcriptomics sections from Control (n=2 individuals) and PVOD (n=1 individual) lung tissues. The two control samples represent the upper and lower halves of the same slide (stitched together). Scale bar=2mm. **b** Spatial mapping of tissue region clusters (Alveoli, Bronchi, Vessel, Unspecified) on spatial transcriptomics spots from Control (left) and PVOD (right) lung samples. **c** Violin plot showing HMOX1 expression levels across tissue regions in Control and PVOD lung samples. *P* values were determined via two-sided Wilcoxon rank-sum test. **d** Violin plots depicting expression of arterial endothelial markers (*KDR, CXCL12*) and venous related marker (*ACKR1*) in vessel regions comparing Control and PVOD groups. *P* values were determined via two-sided Wilcoxon rank-sum test. **e** Violin plots showing arterial and venous endothelial gene set scores in vessel regions of Control versus PVOD samples. *P* values were determined via two-sided Wilcoxon rank-sum test. **f** Volcano plot of differentially expressed genes in vessel regions between PVOD and Control groups. *P* values were determined via two-sided Wilcoxon rank-sum test with Benjamini-Hochberg correction for multiple testing. Significance thresholds were set at |log2 fold change| > 0.5 and adjusted *p*-value < 0.05. The top 5 upregulated and top 5 downregulated genes are annotated in the plot. **g, h** GO biological processes (g) and KEGG pathways (h) significantly enriched (FDR < 0.05) from upregulated genes in PVOD vessel regions. *P* values were calculated using the hypergeometric test with Benjamini-Hochberg correction for multiple testing. Ten relevant terms associated with pulmonary vascular disease are shown, ranked by combined score. Dot size represents the percentage of genes in the gene set, and dot color indicates –log10(FDR). **i** Volcano plot of transcription factor activity differences (z-score normalized AUC scores) between Control and PVOD vessel regions analyzed by limma method. **j** Violin plot showing *ETS1* AUC scores in Control and PVOD vessel regions. **k** Violin plot of *ETS1* expression in venous endothelial cells from scRNA-seq data comparing Control and PVOD groups. *P* values were determined via two-sided Wilcoxon rank-sum test. **l** *ETS1* transcription factor binding motif (metacluster_183.1) obtained from the cisTarget motif collection (v10nr_clust). **m** Spatial distribution of cell type proportions (EC_arterial, EC_venous, Macrophages, Muscular cells, Fibroblasts) inferred by RCTD deconvolution. Color intensity corresponds to the relative abundance of each cell type, with darker colors indicating higher proportions. **n** Heatmaps showing Pearson correlation between RCTD cell type scores and cell death pathway gene set scores in Alveoli (top) and Vessel (bottom) region of the PVOD lung sample (\**P* < 0.05, \*\**P* < 0.01, \*\*\**P* < 0.001). *P* values are indicated in the figures. Source data are provided as a source data file.

Analysis of the vessel regions revealed significant molecular alterations in PVOD. *HMOX1*, a marker of oxidative stress, was markedly elevated in both alveolar and vessel regions of PVOD samples (Fig. 6c). Arterial endothelial markers including *KDR* and *CXCL12* were significantly upregulated, while venous related marker *ACKR1* was reduced in PVOD vessels compared to Controls vessels (Fig. 6d). Gene set scoring confirmed enhanced arterial characteristics with increased arterial scores and decreased venous scores in PVOD vessels (Fig. 6e). Differential expression analysis of vessel regions identified 2,129 upregulated and 590 downregulated genes in PVOD (Fig. 6f). Gene Ontology (GO) and Kyoto Encyclopedia of Genes and Genomes (KEGG) pathway enrichment analysis revealed activation of muscularization and fibrosis-related pathways, including TGF-β and Notch signaling (Fig. 6g, h). Comparison with published PAH spatial transcriptomics data^36^ showed distinct enrichment patterns, with PVOD characterized by prominent angiogenesis, Wnt-β catenin, and Notch signaling (Fig. S16f).

To identify regulatory mechanisms underlying PVOD pathology, transcription factor activity analysis was performed with pySCENIC^37^ analysis. ETS1 emerged as the most significantly upregulated transcription factor in PVOD vessels (Fig. 6i, j), consistent with its elevated expression in venous endothelial cells from sc-RNA seq data (Fig. 6k). cisTarget database analysis confirmed that ETS1 regulates NRP1 and KDR through a conserved binding motif (Fig. 6l).

Cell type deconvolution with matched single cell nuclear RNA sequencing (snRNA-seq) reference data^38^ (Fig. S16g-l) revealed increased venous endothelial cell proportions in PVOD (Fig. 6m). Correlation analysis between cell type scores and cell death pathways showed a specific association between ferroptosis and macrophages in PVOD samples (Fig. 6n), suggesting cell type-specific iron accumulation response in macrophages.

In human lung tissue sections, we also performed multiple immunofluorescence staining for NR2F2 (Veins), CD31 (ECs), and α-SMA (SMCs) to examine the localization of pulmonary venous endothelium. Staining for the arterial markers *NRP1, EFNB2* and *KDR* revealed that these arterial genes were highly expressed on the pulmonary venous endothelium in PVOD patients (Fig. 7a). During vessel formation, EFNB2 regulates arterial formation by upregulating downstream p-ERK, leading to the differentiation of ECs towards the arterial lineage^32, 33^. After treating GCN2-deficient hUVECs with FAC, we observed that NRP1 and EFNB2 protein expression was elevated significantly, accompanied by increased p-ERK activity compared with untreated groups (Fig. 5b). Finally, to confirm the effects of *EIF2AK4* mutation on VEC specification in the context of PVOD, we generated iPSCs from a PVOD patient with *EIF2AK4* mutation and a healthy donor, differentiated the iPSCs into VECs and confirmed NR2F2 and CD31 expression in both lines after mesoderm induction towards VECs (Fig. 7b, d and Fig. S17a). Immunoblotting revealed the elevated expression of p-ERK, ETS1, NRP1 and EFNB2 in PVOD iPSC-VECs compared with Control iPSC-VECs (Fig. 7c, e,f and Fig. S17b). These findings suggest that VECs are prone to differentiate into arterial cells in the context of GCN2 deficiency.

**Fig. 7.**
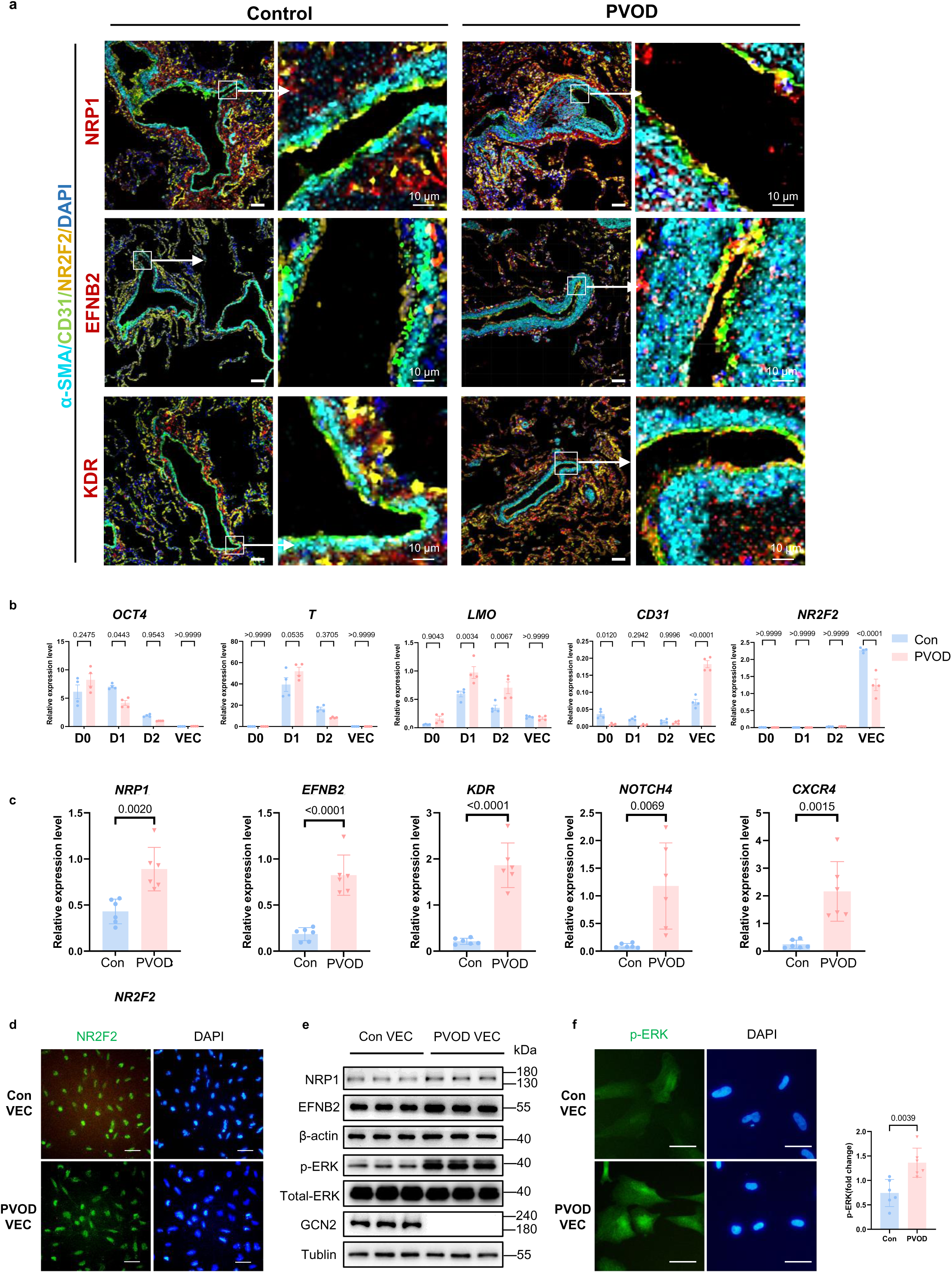
Enhanced NRP1/KDR/EFNB2 expression as a marker set of venous arterialization of the VECs in PVOD patient lung. **a** Representative images of NRP1/KDR/EFNB2 (red), NR2F2 (yellow), CD31 (green), and α-SMA (cyan) immunofluorescence and DAPI (blue) staining of lung tissues from Control and PVOD patients, with the boxed region magnified. Scale bar=50 µm. Enlarged view scale bar=10 µm. The data are representative images. Control, n=7 individuals; PVOD n=9 individuals with similar results. **b** qRT-PCR analyzed the expression levels of representative markers at various stages of VECs differentiation process. Data are presented as the means ± s.e.m. (n=4 replicates); two-way ANOVA with Tukey’s multiple comparison test. *CD31* VEC, *p*=6×10^-12^; *NR2F2* VEC, *p*=4×10^-10^. **c** qRT-PCR analyzed the differential expression levels of candidate genes in Con-iPSC VEC (Con) and PVOD iPSC VEC (PVOD). Data are the mean ± s.e.m., (n=6), unpaired two-sided t-test. *EFNB2* Con vs PVOD, *p*=4.7×10^-4^; *KDR* Con vs PVOD, *p*=8×10^-6^. **d** Immunofluorescence analysis of a venous marker (NR2F2) with DAPI counterstaining. Scale bar=100 µm. The experiment was repeated independently 4 times with similar results. **e** Western blot showing the expression levels of NRP1, EFNB2, p-ERK, Total-ERK in control (Con) or PVOD iPSC-VEC. The experiment was repeated independently 3 times with similar results. **f** Immunofluorescence analysis of p-ERK with DAPI counterstaining. Scale bar=50 µm. Data are the mean ± s.e.m., (n=6 replicates), unpaired two-sided t-test. Each dot represents an individual biological replicate, at least three independent experiments. *P* values are indicated in the figures. Source data are provided as a source data file.

In summary, our findings demonstrated that GCN2 deficiency induces ferroptosis in lung macrophages through the upregulation of HMOX1, which releases iron to induce NRP1/KDR/EFNB2 expression through ETS1 transcription activation in VECs. Activated ERK drives venous arterialization and contributes to PVOD development.

## Discussion

We report here that *EIF2AK4* mutation-induced macrophage ferroptosis contributes to venous arterialization in the pulmonary vasculature and that the inhibition of ferroptosis can reverse pulmonary venous remodelling in animal models of PVOD. This work also shows that mice homozygous for a human disease-causing mutation are susceptible to PVOD with no sex bias, providing a genetic experimental model for this disease and clarifying the molecular characteristics of venous arterialization in human disease.

In this study, we assessed ferroptosis-related gene expression in PVOD patient and model rat lung tissues via scRNA-seq. These observations are consistent with haemosiderin accumulation in macrophages, a pathological characteristic of PVOD^39^. ScRNA-seq analyses revealed that although some upregulation was observed in T cells and NK cells, the highest baseline expression and the most pronounced disease-associated increase occured predominantly in tissue-resident macrophages. Furthermore, GCN2 was reported to control erythrocyte clearance and then affect iron recycling in mouse liver macrophages under stress^30^. The failure to regulate heme and iron by *Eif2ak4* deficient macrophage results in uncontrolled iron release to pulmonary interstitium and bone marrow derived cells (such as dendritic and T cells) infiltration. Increased 4-HNE staining and elevated MDA levels in the lung tissue of PVOD patients and GCN2-deficient mice suggest that the high-iron microenvironment caused by GCN2 deficiency contributes to enhanced lipid peroxidation in the lungs, which may further promote vascular remodelling. PVOD patients have reduced ability to transfer oxygen from inhaled air to the bloodstream, so hypoxia is another critical factor, supporting the study of related phenotypes in our mouse model.

In vitro and in vivo experiments confirmed increased HOMX1 expression in *Eif2ak4*-mutated macrophages^2^, and we also demonstrated upregulation of the transcription factor NRF2 in mutant BMDMs and HT1080 cells. However, HMOX1 inhibition did not reverse this PVOD phenotype. Since PVOD patients experience lung haemorrhage, increased *HMOX1* expression may be required for macrophages to clear extravascular red blood cells^40^. However, our results demonstrated that ferroptosis inhibition not only successfully prevented PVOD development, but also reversed established disease phenotype in *Eif2ak4*^K1488X/K1488X^ mice and MMC rats, suggesting that macrophage ferroptosis inhibition could be an effective treatment strategy for the genetic and nogenetic forms of PVOD.

In this study, we addressed the fate transition from venous lineage to arterial identity in the context of a human disease with various techniques, including scRNA-seq and multiplex immunohistochemistry, and identified NRP1/KDR/EFNB2 as a specific marker set for venous arterialization, consistent with the results of spatial transcriptomic sequencing. Additionally, we demonstrated enhanced ERK phosphorylation in GCN2^-/-^ hUVECs and PVOD-iPSC VECs via immunoblotting and immunofluorescence staining, which is in line with the finding that MAPK activation is triggered by iron treatment^41^. It was also reported that transcription activator ETS1 is the downstream effector of phosphorylated ERK^42^. Our study revealed that activated NRP1/KDR/EFNB2, a core set of markers, can be used for the diagnosis of venous arterialization in venous occlusive diseases and vein stenosis, including predict the outcome and prognosis^43, 44^.

One limitation of the current study is accumulated immune cell in PVOD lungs may contribute to the individually reduced propotion of vascular cell and other cell types. So we sorted CD31^+^ cells by magnetic beads and performed scRNA-seq to obtain transcriptional profiles of pulmonary endothelial cells. To further address this limitation, we employed spatial transcriptomics and Single-nuclear transcriptomics to better distinguish and analyse EC types and their expression profiles, and provide a more balanced perspective on the cellular landscape of the affected lung tissue. Another limitation is inadequate comparison between PVOD and other forms of pulmonary arterial hypertension (PAH). Although we conducted a comparative analysis using scRNA-seq data from three idiopathic PAH (IPAH) lung tissue samples alongside our PVOD dataset (Fig. S18), as well as an additional comparison with published PAH spatial transcriptomics data (Fig. S16f), our findings revealed both shared and distinct molecular features. However, a comprehensive understanding of the unique molecular signatures distinguishing PVOD from PAH remains to be fully elucidated. Such insights are important for clarifying the distinct pathophysiology of PVOD and reinforcing its clinical differentiation from PAH. In future studies, screening for the relevant disease types using this venous arterialization marker set to ensure that targeted clinical trials involving ferroptosis inhibition will be crucial.

Overall, we provide a genetic model to study venous arterialization in human disease. Our study builds on previous reports by clarifying the details of the interaction between *EIF2AK4* defects and iron metabolic alterations in the pulmonary vascular niche, suggesting that NRP1/KDR/EFNB2 could be hallmarks of pathogenic venous arterialization and that targeting the ferroptosis pathway is worth investigating for the treatment of PVOD and other related vascular diseases.

## Methods

### Collection of Pulmonary Veno-Occlusive Disease (PVOD) lung tissue

Lung tissue samples from PVOD patients(with no sex bias) were obtained during lung transplantation procedures after written informed consent was obtained from all patients. Informed consents were obtained from all human subjects. All procedures were approved by the Institutional Review Boards (IRBs) of the Second Affiliated Hospital of Zhejiang University School of Medicine (IRB-2021-588 and IRB-2024-1507) and Wuxi People’s Hospital NO. (2015)36. The diagnosis of PVOD was confirmed via exome sequencing, which identified biallelic mutations in the *EIF2AK4* gene. The samples included multiple sections of proximal, middle, and distal lung tissue, each approximately 2 cm in size. Human lung tissues were obtained from PVOD and unused donor- explanted lungs as controls. The data are shown in Supplementary Table 1.

### Single-Cell RNA Sequencing and Analysis

Fresh lung tissues from three PVOD patients (one processed with CD31^+^ magnetic bead enrichment) and whole lungs from MMC-treated and control rats (n = 2 each) were enzymatically dissociated into single-cell suspensions for single-cell RNA sequencing (scRNA-seq). Additionally, a frozen lung tissue sample from one PVOD patient was processed for single-nucleus RNA sequencing (snRNA-seq) to be paired with subsequent spatial transcriptomic analysis. Following quality control (> 90% viability for cells, > 70% intact nuclei for frozen tissue), samples were processed using 10x Chromium 3’ v3 chemistry and sequenced on Illumina HiSeq 2000.

Raw data were aligned using Cell Ranger v6.0.0 to GRCh38 (human) or Rnor 6.0 (rat) reference genomes, with pre-mRNA reference included for snRNA-seq to capture intronic reads. The entire analysis workflow combined the use of Seurat (v4.3.0), Scanpy^45^ (v1.11.2), and OmicVerse^46^ (v1.7.1). Quality control retained cells/nuclei with < 15% mitochondrial content (< 5% for nuclei) and sample-specific gene detection thresholds. Doublets were removed using Scrublet^47^ (v0.2.3). After normalization and scaling, principal component analysis was performed on 3,000 highly variable genes. Batch effects were corrected using OmicVerse. single. batch_correction (scVI or Harmony parameters). Cell types were annotated with assistance from CellTypist^48^ (v1.7.0) using the Human Lung Cell Atlas (HLCA)^23^ reference, followed by graph-based clustering via the Leiden algorithm. Differentially expressed genes were identified using scanpy.tl.rank_genes_groups (Wilcoxon test) with adjusted p-value < 0.05 and log2 fold change > 0.5.

Gene set enrichment analysis (GSEA) was performed on differentially expressed genes between Control and PVOD groups within each cell type subpopulation. Single-sample gene set enrichment analysis (ssGSEA) was implemented using the "escape" package (v2.2.3) for gene set scoring across individual cells, incorporating ferroptosis gene sets from FerrDb^27^ and the iron accumulation signature (IAS) gene set from Maus et al.^26^ to assess iron-related cellular states. Cell-cell communication networks were inferred using CellChat^49^ (v1.6.1) to predict ligand-receptor interactions among immune and endothelial cell populations.

### 10x Genomics Visium Spatial Transcriptomics and Analysis

Formalin-fixed paraffin-embedded (FFPE) tissue sections (5 μm thickness) from one PVOD patient and two healthy control lungs were processed using the Visium Spatial Gene Expression for FFPE Kit (10X Genomics) following manufacturer’s protocols. Both control samples were captured on a single chip. Sections were deparaffinized with xylene, rehydrated through graded ethanol series, and subjected to H&E staining for morphological assessment. Following probe hybridization and ligation-based target enrichment specific to FFPE workflow, libraries were sequenced on the Illumina NovaSeq platform. Raw FASTQ files and histology images were processed using Space Ranger v2.0.0 (10X Genomics) to generate gene-spot matrices aligned to GRCh38.

Spatial data integration across the three samples (control1, control2, PVOD1) was performed using STAligner^35^. Normalized data underwent principal component analysis, and graph-based clustering was performed using the Leiden algorithm. Arterial and venous signatures were scored using scanpy.tl.score_genes. Differentially expressed genes between control and PVOD groups were identified using scanpy.tl.rank_genes_groups (Wilcoxon rank-sum test) with adjusted p-value < 0.05 and log2 fold change > 0.5. Pathway enrichment analysis was performed using gseapy (v1.1.9) for GO, KEGG, and Hallmark gene sets.

Transcription factor regulatory network analysis was conducted on vessel regions using pySCENIC^37^ (v0.12.1). Cell type deconvolution was performed using RCTD^50^ (spacexr v2.2.1), with the paired snRNA-seq data from the same PVOD patient serving as reference for the PVOD spatial data, and publicly available healthy lung snRNA-seq data^38^ for the control spatial data. For each spot, cell type proportions were correlated with gene set scores for four cell death pathways (ferroptosis, apoptosis, necroptosis, pyroptosis) using Pearson correlation. The predominant cell type per spot was defined as "maxtype" based on highest deconvolution score.

### MMC-Induced rat model and treatments

All the rat experiments performed in the present study were approved by the Animal Institutional Review Boards (AIRB) of the Second Affiliated Hospital of the Zhejiang University School of Medicine (AIRB-2021-967 and AIRB-2024-327). Five-week-old female Wistar rats (150‒200 g) were randomly assigned to treatment groups. The control rats were given intraperitoneal injections of a control solution (50% PEG300 (MCE, HY-Y0873) + 50% saline), whereas the MMC rats received MMC (3 mg/kg, once a week for two weeks, MCE, HY-13316). Fer-1 (1 mg/kg, Selleck, S7243) was administered intraperitoneally every other day, with the control group receiving saline. For the prevention model, saline or Fer-1 was given beginning on the first day of MMC treatment for 35 days. For the therapeutic model, treatment started on day 21 post-MMC treatment and continued every other day for 14 days. The HO-1 agonist hemin (25 mg/kg, Sigma, H9039) and the HO-1 inhibitor Znpp (10 mg/kg, MCE, HY-101193) were administered intraperitoneally every other day for 35 days, starting from the first day of MMC treatment, with the control groups receiving saline or 50% PEG300 + 50% saline, respectively. At the time of sacrifice, the rats were anaesthetized with 40 mg/kg sodium pentobarbital and inhaling in 0.5% isoflurane, the anaesthesia depth was monitored with pedal reflex.

### Generation of experimental PVOD mice and treatments

All the mice experiments performed in the present study were approved by the Animal Institutional Review Boards (AIRB) of the Second Affiliated Hospital of the Zhejiang University School of Medicine (AIRB-2021-967 and AIRB-2024-327). A guide RNA (gRNA) targeting exon 33 of *Eif2ak4* was designed with the sequence 5′-GCAAACAGAAAGCGTGTATTGG-3′ for use in CRISPR/Cas9 (Shanghai Model Organisms). The C57BL/6J mice with the mutation c.4413- 4416del were identified and bred to obtain homozygous mutants (*Eif2ak4*^K1488X/K1488X^). 4-month-old C57BL/6J mice (25–30 g, equal male-to-female ratio) were assigned to the following groups: the WT with or without hypoxia, *Eif2ak4*^K1488X/K1488X^ with or without hypoxia. Hypoxia treatment (10% oxygen) for 6 weeks, and then return to normoxia for 1 week. Fer-1 (1 mg/kg) was administered intraperitoneally every other day; in the prevention model, Fer-1 was administered from the first day of hypoxia for 42 days, and in the therapeutic model, Fer-1 was administered starting on day 21 for 21 days. Hemin (25 mg/kg) and Znpp (10 mg/kg) were administered intraperitoneally every other day, with the control groups receiving saline. Mice were housed under standard conditions in pathogen-free, individually ventilated microisolator cages in a room with a 12 h light/dark cycle, and a temperature- and humidity-controlled environment of 20-26°C and 30- 70% humidity, with access to standard laboratory chow diet and water ad libitum. At the time of sacrifice, the mice were anaesthetized with 0.15 mL of 1% sodium pentobarbital and inhaling in 0.5% isoflurane, the anaesthesia depth was monitored with pedal reflex.

### Bone marrow chimera mouse model

Bone marrow cells were isolated from the femur bone cavities of WT and *Eif2ak4*^K1488X/K1488X^ mice, transplanted into 7-Gy irradiated WT recipient mice via tail vein injection. Blood was collected three weeks after bone marrow transplantation for DNA isolation with a commercially available DNA isolation kit (Tiangen, DP304) and the *Eif2ak4* transgene was detected by PCR using the genotyping primers of *Eif2ak4*^K1488X/K1488X^ mice to detect the transgene. The chimeric recipient mice were subjected to hypoxia 3 weeks after transplantation.

### Intratracheal administration of iron to mice

Ferric ammonium citrate (FAC, 50 mM, F3388, Sigma) was administered intratracheally to 3- month-old C57BL/6J WT and *Eif2ak4*^K1488X/K1488X^ mice^26^. The mice were anaesthetized with 0.15 mL of 1% sodium pentobarbital and inhaling in 0.5% isoflurane, and 10 μL of PBS/FAC was delivered intratracheally weekly for two weeks. Pulmonary function, the right ventricular systolic pressure (RVSP), right ventricular hypertrophy (RVH), and lung histology were evaluated four weeks after treatment initiation.

### Hemodynamic measurements in rats and mice

Identical methods were employed for haemodynamic measurements in both the rats and the mice. After the animals were anaesthetized, a millar catheter (mice PE-10, rats PE-90, Smith Medical) was inserted into the right ventricle via the right jugular vein to measure the RVSP. Continuous signals were collected using a physiological monitor for data analysis. To assess the RVH index, the hearts were dissected, and the right ventricle was isolated, washed with PBS, and weighed. The RVH index was calculated with the following formula: RVH index = (right ventricular weight/(left ventricle + septum weight)) × 100% (1). The total pulmonary vascular resistance index (TPVRI) was calculated using the following formula: TPVRI = RVSP (mmHg)/CI (2), where CI is RVCO (mL/min) * 100/body weight (g) (3). The RVSP was measured via right heart catheterization, and the RVCO was estimated via echocardiography. For a statistical analysis of vascular remodelling, immunohistochemical staining of α-SMA was performed, and images were analysed using the VS200 system. Rat vessels (70–250 μm) were analysed for lumen-to-total area ratios and media layer thickness, respectively. Microvessels were classified on the basis of α-SMA expression levels.

### Immunohistochemical Staining

The paraffin-embedded sections were dewaxed, hydrated, and subjected to antigen retrieval. Endogenous peroxidase activity was quenched, and the sections were incubated with primary antibodies (against GCN2, 4-HNE, α-SMA, and HMOX1) overnight. Following secondary antibody incubation and DAB chromogen development, the sections were counterstained with haematoxylin, dehydrated, and mounted. Quantification of IHC images was performed in a blinded manner using ImageJ (Fiji; https://imagej.nih.gov/ij/). Specifically, the intensity of staining was scored, and the percent of postively stained areas was calculated for analysis. Antibodies used for immunostaining are listed in Supplementary Table 4.

### Classification of arteries and veins

Arterial vessels accompany bronchial structures, demonstrating distinct internal and external elastic laminae with densely arranged smooth muscle cells in the tunica media.Venous vessels exhibit only an external elastic lamina, featuring thinner vessel walls with loose connective tissue and significantly larger luminal diameters compared to corresponding arteries. Immunofluorescence was applied for venous markers (NR2F2/α-SMA/CD31) and artery markers (α-SMA/CD31) showing different types of vessels in the lungs.

### Percent wall thickness of artery and vein

After fixing in 4% paraformaldehyde solution, embedding and sectioning, the 5 µm slides of mouse pulmonary vessels (50–150 μm) were stained with α-smooth muscle actin to access the medial thickness^51, 52^. Double-blind analysis of sections was completed by light microscopy (OLYMPUS VS120). The external vessel perimeter (measuring from outside margin of external α-smooth muscle actin lamina), and internal vessel perimeter (inside margin of internal α-smooth muscle actin lamina) were measured using (OlyVIA). 15 images of 50-100 µm vessels at 40× fields were acquired randomly from 5∼7 mice in each group. Radius=perimeter/π/2 (4), and medial thickness=external radius-internal radius (5). Standardized the thicknesses by using the ratio of thickness to diameter, percent wall thickness=2*medial thickness/external diameter (6)^53^.

### Prussian Blue staining with DAB enhancement

The paraffin-embedded sections were dewaxed and hydrated. The sections were then stained with 2% potassium ferrocyanide and 2% hydrochloric acid for 30 minutes, followed by three washes with distilled water. The chromogen DAB was applied for 5‒10 minutes. The sections were counterstained with haematoxylin for 1 min, blued in PBS and rinsed with tap water. The sections were subsequently dehydrated in absolute ethanol, cleared in xylene, and mounted with neutral balsam.

### Heme quantification assay

Heme was quantified using the Heme Iron Assay Kit (Abcam, ab272534). Tissue samples (0.1 g) were homogenized in 1 mL of PBS and centrifuged at 13500 x g for 10 minutes, after which the supernatants were collected. The assay wells received 50 µL of water (blank), the calibration solution, or a sample, followed by 200 µL of deionized water (blank/calibration wells) or reaction reagent (sample wells). The absorbance at 400 nm was measured after a 5-minute incubation period. The heme concentration was calculated as follows:

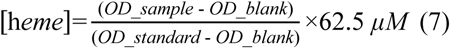

### Measurement of tissue non-heme Iron

Frozen lung tissue was sectioned, weighed, and digested in NHI acid at 65–70°C for ≥72 hours. The homogenized tissue was centrifuged at 13500 x g for 10 minutes. The supernatant was incubated with equal volumes of BAT buffer and a standard iron solution at room temperature for 10 minutes. The absorbance was read at 535 nm, after which the iron concentration was determined using a standard curve; the iron concentration is expressed as µg of iron per g of wet tissue.

### Lipid peroxidation (MDA) Assay

Lipid peroxidation was measured with an MDA Assay Kit (Biyuntian, S0131S). Frozen lung tissue (0.1 g) that had been homogenized in 1 mL of PBS was centrifuged at 13500 x g for 10 minutes. An MDA working solution was prepared and mixed with the lung homogenate, a standard, or PBS, followed by heating in a boiling water bath for 15 minutes. The samples were cooled and centrifuged, and the absorbance was measured at 532 nm and 450 nm.

**Isolation and culture of mice primary Bone Marrow-Derived Macrophages (BMDMs)** Femurs and tibias from euthanized mice were flushed with 1640 medium, and bone marrow cells were isolated (half male and half female). After red blood cell lysis, the cells were resuspended in differentiation types of medium (1640 medium, 10% serum, 15% L929 cell-conditioned medium) and plated. Differentiation was maintained by changing the medium every other day until day 7.

BMDMs were harvested via 0.25% trypsin. For the cell viability and ferroptosis assays, BMDMs were treated with FAC or RSL3, and the staining assays were performed with BODIPY581/591^54^.

### Treatment of endothelial cells with macrophage supernatants

BMDMs differentiated 7 days were treated with 300 µM FAC for 24 hours, and the supernatants were collected. The cell supernatant was filtered through a 0.22 μm filter and treated with endothelial cells for 72 hours.

### Preparation of lentivirus vectors

The gRNA targeting mouse exon 33 was designed at http://crispor.tefor.net/. The gRNA sequence was 5′-GCAGACAGAGAAGCGTGTGC-3′. This gRNA was subsequently cloned and inserted into pLentiCRISPRv2 and transfected into 293T cells along with the packaging vector pSPAX2 and the envelope vector VSVG. The supernatants were collected 48 hours post-transfection, filtered, and concentrated. The viral pellet was resuspended in the target cell culture medium for subsequent infection.

### Cultivation and treatment of endothelial cells and HT1080 cells

Human umbilical vein endothelial cells (hUVECs, CBP60340), human pulmonary artery endothelial cells (hPAECs, Lonza, CC2530), and human pulmonary microvascular cells (hPMECs, Oligobio, oligo876L) were cultured in ECM medium with 5% FBS. Lentivirus were used for GCN2 knockdown, which was confirmed by Western blotting. Endothelial cells were preconditioned in low-serum medium before treatment with FAC (Sigma, F3388), TNF (Abclonal, RP00993), GDF15 (R&D, 8146-GD-025), or CCL3 (Abclonal, RP01628). The cells were harvested for protein or RNA analysis following treatment. HT1080 cells (CBP60250) transfected with lentivirus and selected with puromycin were propagated in high-glucose medium supplemented with 10% FBS. For ferroptosis pathway detection, the cells were treated with FAC, and subsequent analyses included protein extraction, Per’s DAB iron staining, and calcein assays.

### Protein extraction and quantification and Western blotting

Protein extraction was performed using the Pierce™ BCA Protein Assay Kit (Thermo, 23227). Adherent cells and tissue samples were lysed and homogenized in RIPA buffer. The supernatants were mixed with loading buffer, boiled, and stored at -80°C (long-term). Protein quantification was performed with a BCA working solution and measuring the absorbance at 562 nm after 30 minutes at 37°C. For Western blotting, samples were separated by SDS‒PAGE gels and then transferred to PVDF membranes with standard operation. The membranes were blocked with QuickBlock™ Western Blocking Buffer, and incubated overnight at 4°C with primary antibodies and followed with secondary antibody incubation at room temperature. Detection was performed using enhanced chemiluminescence (ECL, 1705060, Bio-rad) reagent. Source data are provided as a source data file. All antibodies in this study were commercially purchased and have been validated by the vendors for species and application. Validation data are available from the respective vendor’s respective websites. Antibodies for western blotting are listed in Supplementary Table 3.

### RNA extraction, Reverse Transcription, and Quantitative Real-Time PCR (qRT-PCR)

Total RNA was extracted using TRIzol reagent (Genstar, P118). After mixing lysate with chloroform, extracted RNA was incubated at -20°C overnight. Purified RNA was dissolved in RNase-free water. Reverse transcription was carried with gDNA Clean Reaction Mix and M-MLV reverse transcriptase with standard protocol (Accurate biology, AG11728). Quantitative Real-time PCR was performed using ChamQ Blue Universal SYBR qPCR Master Mix (Vazyme, Q312) on a Roche Light Cycler 480 instrument. The data were analysed using Light Cycler 480 software. Primer sequences used are listed In Supplementary Table 2.

### Transwell endothelial cells and smooth muscle cells Coculture Assay

To investigate the effects of ECs on SMC proliferation and migration, 0.4 μm and 8 μm Transwell chambers (Falcon, 353095 and 353097) were used, respectively. hUVECs were seeded in 24-well plates, starved in high-glucose medium supplemented with 0.2% FBS for 24 hours, and then treated with 300 μM FAC for 72 hours. SMCs were seeded in Transwell chambers with high-glucose medium supplemented with 10% FBS and allowed to attach, after which the medium was replaced with medium containing 0.2% FBS. Chambers were placed in the wells with the treated hUVECs for 24 hours of coculture. SMCs were fixed with PFA, stained with DAPI, and counted under a microscope.

### Three-dimensional Coculture of endothelial cells and smooth muscle cells

GFP-tagged hUVECs and red-stained SMCs were cocultured to form a 3D system. GFP-positive hUVECs were selected and expanded. SMCs were stained with PKH26. A mixture of hUVECs and SMCs was combined with ECM and Matrigel (Corning, 356234), and the mixture was then dispensed into μ-Slide chambers (Ibidi, 81506). After incubation, medium supplemented with 0.2% FBS and 660 μM FAC was added, and the medium was replenished daily. Images were captured using a confocal microscope, and the vascular network was analysed via ImageJ software.

### Multicolour immunofluorescence staining

The tissue sections were deparaffinized, subjected to antigen retrieval with EDTA solution, and blocked with 3% BSA. Primary antibodies were applied overnight at 4°C, followed by incubation with HRP-conjugated secondary antibodies. TSA staining (servicebio, G1255) was performed sequentially with multiple antibodies. The sections were stained with DAPI and treated with an autofluorescence quencher. Images were acquired using a confocal fluorescence microscope.

### Calcein Staining

Primary mice BMDMs and HT1080 cells were seeded in a 12-well plate and treated with 100 μM FAC for 24 hours. The cells were stained with calcein AM (Beyotime, S2012), incubated in the dark at 37°C for 10 minutes, and washed with PBS. The fluorescence intensity was measured using a microplate reader at an excitation wavelength of 494 nm and an emission wavelength of 524 nm.

### GSH/GSSG Assay

Total and oxidized glutathione levels were measured using a GSH/GSSG Assay Kit (Beyotime, S0053). The tissue samples were homogenized and centrifuged, and the supernatant was used for the analysis. The samples were treated to remove GSH, and the absorbance at an OD of 412 nm was measured using prepared standards and reagents.

### ELISAs

ELISAs for TNF, GDF15, and CCL3 were conducted using specific kits (ABclonal, Human TNF, RK00030; Mouse TNF, RK00027; Human GDF15, RK00086; Mouse GDF15, RK00369; Mouse CCL3, RK04218). The tissue samples were homogenized, and the supernatants were collected. Standard curves were prepared, samples and standards were incubated with biotinylated antibodies and streptavidin-HRP, and the absorbance was measured at OD450 nm with a reference at OD630 nm using a microplate reader.

### Differentiation of Venous endothelial cells

iPSCs were generated from PBMCs of a healthy donor (male) and PVOD patient (male) following established protocols^55^. When differentiation was initiated, iPSCs were dissociated into single cells using TrypLE Express (Thermo Fisher, 12604013) and plated onto Matrigel-coated plates at a density of 25,000-50,000 cells/cm². Freshly seeded iPSCs were allowed to adhere and recover for 24 hours in mTeSR medium supplemented with 1 mM thiazovivin (Sigma, SML1045) before initiating differentiation. Differentiation into venous endothelial cells was conducted in Chemically Defined Medium 2 (CDM2), the medium was sterilely filtered through a 0.22μm filter prior to use. Media were exchanged at 24-hour intervals with CDM2 basal medium supplemented with various cytokines and chemicals. After 4 days, we yielded vein endothelial cells (VECs)^56^.Then, iPSC-derived VECs were enriched for CD31^+^ cells using CD31 microbead-based Magnetically Activated Cell Sorting (MACS, Miltenyi Biotec, 130-091-935).

### Statistics and Reproducibility

Statistical analysis was conducted using GraphPad Prism v9.0.0 or R software v4.2.3. The figure legends or Methods sections detail the statistical tests conducted and the repeat numbers. *P* value < 0.05 was considered statistically significant unless otherwise stated.

### Data availability

Access to the human data is controlled and requires data access committee approval. Source data are provided with this paper.

### Code availability

All analyses were conducted using existing software packages detailed in the Methods. No custom code was generated.

## Acknowledgements

We thank Prof. Zhou Zhou for kindly giving two PVOD patient iPSCs lines and Prof. Xiaofang Fan for catheterization of mice. We also thank Dr. Zhang Ruifeng for clinical consultation and Dr. Cai zongye for providing oxygen sensor on hypoxia chamber, Dr. Yu Zhou and Man Huang of Second Affiliated Hospital of Zhejiang University School of Medicine provided the patient sample storage and delivery. Dr. Dan Liu and Jing Feng from Tianjin General Hospital provided collaboration based clinical immunostaining training. We also thank core facilities (Zhejiang University School of Medicine) providing the confocal microscopy and other instruments and Novel Bio Co., Ltd for transcriptomic analysis. This project received funding from the National Key Research and Development Program of China (2023YFC2507100 to J.Y.C., 2023YFC2507103 to J.Y., 2021YFA1100500 to X.X. and J.Y.), the National Nature Science Foundation of China (82470431 to J.Y., 81670054 to J.Y., 82100364 to J.Y.Z.), the China Postdoctoral Science Foundation (2022M712751 to J.Y.Z.). This preprint has not undergone any post-submission improvements or corrections. The Version of Record of this article is published in Nature Communications, and is available online at https://doi.org/10.1038/s41467-025-64035-4

## Author contributions

J.Y., design of the research studies, revision and finalization of the manuscript. J.Y.Z., conception, design, acquisition, analysis and interpretation of data and to the drafting of the article. P.M., help with design and the analysis of experiments related to iPSC vein endothelial differentiation. T.F.Z., help with the analysis of sc-RNA seq and 10× Visium transcriptome. B.Q.Y., providing lung tissue samples of PVOD patients. Y.H.Q., providing patient information. Y.N.L., help with the analysis of data. K.J.Y., providing advice. J.Y.C., the major contribution on providing the PVOD patient samples. F.D.W., providing supervision on iron metabolism study. All authors contributed to revision of the article.

